# Effects of Long-Term Supplementation of Eggs on Growth, Biochemical Indices, and Microbiota of Rural Thai Primary School Children

**DOI:** 10.1101/2022.08.17.22278880

**Authors:** Sophida Suta, Apinya Surawit, Pichanun Mongkolsucharitkul, Bonggochpass Pinsawas, Thamonwan Manosan, Suphawan Ophakas, Tanyaporn Pongkunakorn, Sureeporn Pumeiam, Kitti Sranacharoenpong, Sawannee Sutheeworapong, Patcha Phuangsombut, Sakda Khoomrung, Iyarit Thaipisuttikul, Korapat Mayurasakorn

## Abstract

**Summary:** *Background:* Protein-energy malnutrition is still problematic worldwide. It directly impacts growth and development, especially in children. We investigated the long-term effects of egg supplementation on the growth, biochemical indices, and microbiota of primary school children.

*Methods:* A randomized controlled cluster study was carried out in six rural schools in Thailand. Participants were randomly assigned into three groups: 1) whole egg (WE) – consuming 10 additional eggs/week [n = 238], 2) protein substitute (PS) - consuming yolk-free egg substitute equivalent to 10 eggs/week [n = 200], and 3) control group (n= 197]). Demographic and biochemical indices, and microbiota composition were measured at weeks 0, 14, and 35.

*Findings:* 635 students (8 to 14 years old) were recruited (51·5% female). At baseline, 17% of the participants were underweight, 18% were stunted, and 13% were wasted. At week 35, compared to the control group, body weight and height increased significantly in WE (3·6 ± 23·5 kg, *P*<0·001 and 5·1 ± 23·2 cm, *P*<0·001). No significant differences in weight or height were observed between PS and Control. Prealbumin levels were higher (1·5 ± 8·158·6314 mg/dL, *P*<0·001) in WE, but not in PS, compared to control. Significant decreases in total cholesterol, triglycerides, and LDL cholesterol were observed in the WE, but not in the PS groups. HDL cholesterol tended to increase in WE (0·7 ± 25·2 mg/dL, *ns*). Neither the alpha nor beta diversity of the bacterial diversity was significantly different among all groups. After WE supplementation, the overall relative abundance of *Bifidobacterium* increased by 1·28-fold as compared to baseline and the differential abundance analysis also indicated that *Lachnospira* increased significantly and *Varibaculum* decreased.

*Interpretation:* Long-term whole egg supplementation is an effective, feasible and low-cost intervention to reduce protein-energy malnutrition, particularly in low-middle-income countries. Whole egg supplementation improves growth and nutritional biomarkers, and positively impacts gut microbiota without adverse effects on blood cholesterol levels.

*Funding:* Agricultural Research Development Agency (ARDA) of Thailand (PRP6105022310, PRP6505030460).

## Introduction

Protein-energy malnutrition (PEM) is still a nutritional problem worldwide that has a lifelong repercussion on schoolchildren’s growth and development.^1,2^ The COVID-19 pandemic has even worsened essential nutritional status, leading to unprecedented health problems, especially in developing countries.^3^ Lack of sufficient amount or quality of protein leads to poor diet, poor school performance, and affects social and emotional development, particularly among vulnerable groups and children under years of age.^4,5^ Intake of proteins below physiological needs results in reduced growth and an immune system that is susceptible to disease, infection, and mortality in early life.^3^ According to World Health Organization data from 2019, 24·7% of children in Southeast Asian countries were malnourished^6^, many of whom lived in households with insecure income. These stunted children were more likely to have a below-average Intelligence Quotient.^7^ School closures led to the disruption of the free school lunch program, exposing millions of children to food insecurity.^4^ Our preliminary survey in 2021 showed financial difficulties caused by the lockdown forced families to choose much cheaper and low-quality food choices, exacerbating severe malnutrition and disparity in many societies. This leads to an increased risk of obesity, stunting, and underweight.

Malnutrition is a complex public health problem arising from economic hardship and other environmental and biological factors.^8^ Many countries, including Thailand, continue to beautifully tackle the malnutrition crisis by implementing effective interventions such as school lunch programs, cash transfers, special supplemental nutrition programs, proactive nutrition education and fortified foods.^3,9^ In Thailand, the government has provided free lunch and a serving of milk every school day for primary school children since 1993 and malnutrition has improved over time.^10^ Since 2013, Thai School Lunch, an online platform, has been released to ease school preparation of lunch menus.^11^ Still, the 5th National Health Examination Survey of Thailand in 2014 showed that about 400 000 (3·5%) Thai children aged 1 to 14 years were stunted, while 470 000 (4·1%) children were still underweight. On the contrary, the prevalence of overnutrition in children has increased and is associated with the early onset of non-communicable adult chronic diseases.^12^ This can be caused by imbalanced macronutrients and micronutrient intake, particularly vitamin A, iron, vitamin D, and calcium.^10,13^ Eggs are a common food worldwide that provides approximately 150 kcal/100 g, >50% of adequate intake of these critical micronutrients, high-quality protein, culinary versatility, and are more affordable than other animal-source foods.^14^ Additionally, there are no major religions that place restrictions on egg consumption and eggs are one of the eighth food groups used for better nutritional feeding indicators.^15,16^ Eggs are a rich source of cholesterol and have high concentrations of choline^17^, which plays an integral role in neurotransmitters, cell membrane signaling, methyl metabolism, lipid transport, and metabolism.^18-20^ Many studies demonstrated that whole egg consumption results in increased blood protein and lipoprotein levels.^21-23^ Recent evidence from a middle-income country suggests the early introduction of 1 medium size egg per day for 6 months markedly enhanced growth in young children. ^8,24^ Even though these results seem to be quite beneficial, further long-term intervention studies are necessary to fully understand the synthesis.

Numerous studies have shown that a healthy gut microbiota is crucial for the interaction between food intake and host health.^25-27^ The microbiota participate in extensive molecular crosstalk with the host (triggered by small signaling molecules and immune modulators, *etc*.), influencing nutrient metabolism, the immune system, mood, sleep cognition, and memory.^28-30^ Malnutrition has been linked to gut dysbiosis, a disproportionate composition of the microbiota^28^ by altering healthy and pathogenic microbiota that efficiently process foods or produce vitamins. These changes are capable of impacting the healthy mucosal immune system. Environmental factors, such as diet, stress, chemical exposures, antibiotic administration and geographic location, are linked to changes in the composition of the gut microbiota and can contribute to, or exacerbate, a variety of disorders.^31^ Alterations in the composition of the gut microbiota have been observed in cardiovascular disease (CVD), including atherosclerosis, hypertension, malnutrition, and heart failure.^32^ For example, the number of species in the phylum *Proteobacteria* increases in malnourished infants, while the number of species in the phyla *Bifidobacterium* and *Lactobacillus* decreases.^33^ Recent short-term studies revealed that egg consumption improved hemostasis of intestinal flora and gut microbial function, despite the absence of relative changes in taxonomic abundances and alpha and beta diversities.^34,35^ Consumption of two to three eggs per day increased plasma choline and betaine levels in healthy postmenopausal women and subjects with metabolic syndrome^36^, while inflammatory, metabolic, and oxidative stress markers were not altered.^34^ Therefore, egg consumption may help not only to address malnutrition, but may also ameliorate problems with vascular and intestinal function related to alterations in the gut microbiota.^37^

While the short-term benefits of egg supplementation may have been demonstrated, there is considerable controversy regarding its long-term consequences. Therefore, we investigated the long-term effects of egg supplementation on growth, blood biochemical indices, and gut microbiome in Thai primary school children enrolled in a school lunch program. Blood cholesterol and gut microbiota diversity were also determined.

## Methods

The study protocol was approved by the Institutional Review Board of Siriraj Hospital, Mahidol University (COA No. Si 322/2017). This clinical trial was registered to Clinicaltrials.gov (Protocol NCT04896996). The volunteers’ required written informed consent was obtained from the parents or legal guardians of the participating children prior to starting the study. Their identities are protected. The trial profile is presented in **Figure 1**.

**Figure 1:**
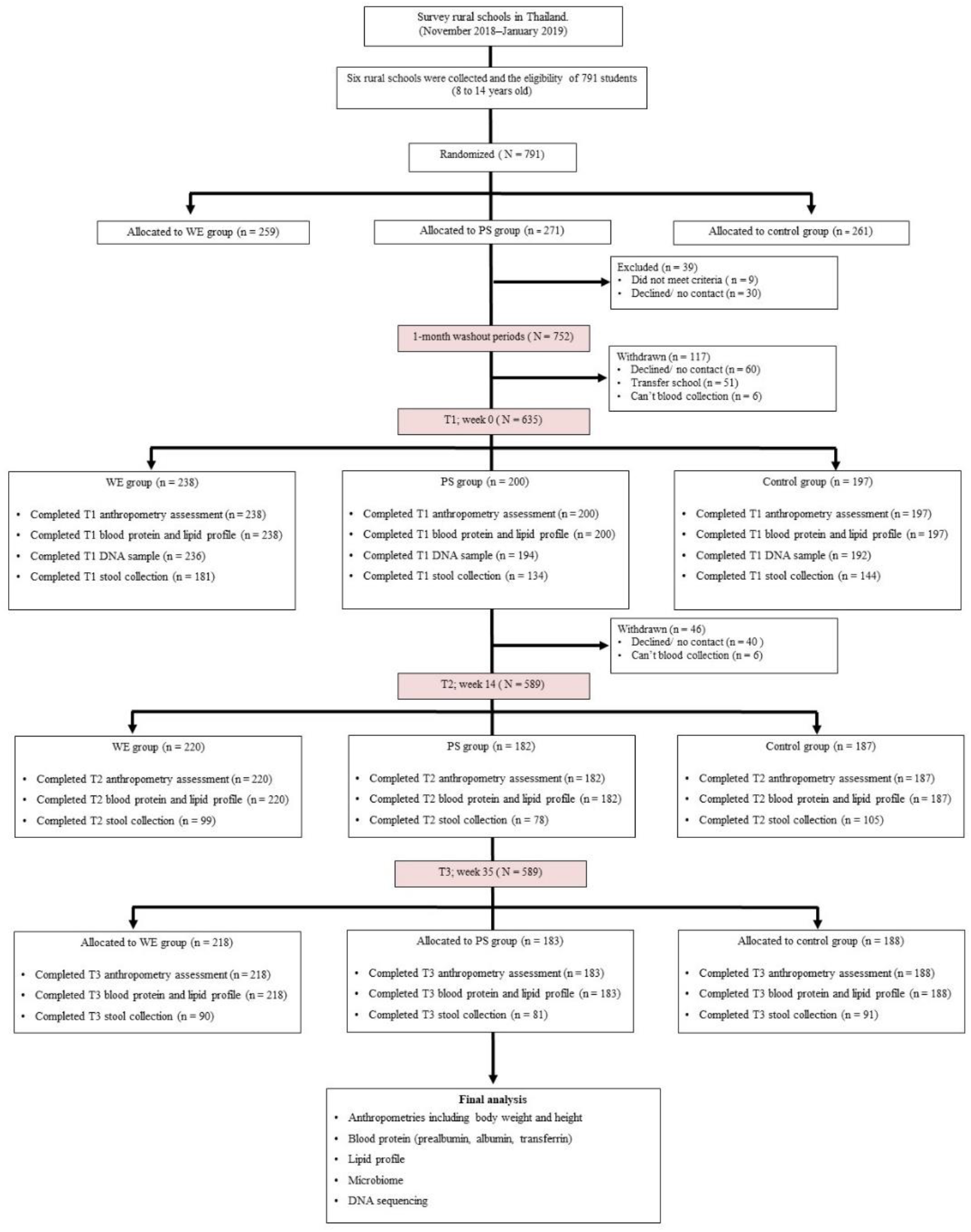
Trial profile. PS= protein substitute group. WE= whole egg group.

### Study population

A cluster randomized controlled trial with parallel design was performed at six rural schools in Nakhon Pathom (Central), Chachoengsao, Chon Buri (Eastern), and Ratchaburi (Western) in Thailand. This was aimed at evaluating the superiority of long-term additional egg consumption of children’s growth and biochemical parameters. This was a geographical distribution pattern of diets, and all were about two to three hours’ drive from Bangkok. This study was aimed at rural schools. The school locations were considered rural areas according to a low population density and no franchise convenience stores within a 10-kilometer radius. We chose rural schools where >10% of all students were underweight based on weight-for-age (W/A) measurements. Primary school participants aged 8 to 14 years were recruited between May 2019 and March 2020. Children with egg allergy were excluded.

### Study Design

All participants from each school were recruited and randomly assigned to three groups based on weight-for-age criteria to ensure that all groups were homogeneous:1) whole egg (WE) - consumed 10 additional whole chicken eggs/week, 2) protein substitute (PS) - consumed a yolk-free egg substitute equivalent to 10 eggs/week, and 3) control group. Each classroom in each school was assigned to a group in one of the three groups to reduce group confusion and maintain group compliance. We evaluated the general effects of the intervention on the outcome but did not perform stratified analyses. All participating schoolteachers received a training session on school lunch menus and were responsible for monitoring the participants throughout the study. All six schools were asked to prepare the same school lunch menus if possible to standardize the calories and nutritional composition of meals according to the national school lunch program.^10^

Before conducting the intervention, all participants were asked to maintain their usual consumption of eggs and dietary cholesterol for four weeks (washout period [week-4). Participants who were randomized to an intervention (WE and PS) continued their usual dietary habits. The intervention was delivered individually to each classroom at their general lunch time. The WE group received a cycle, ready-to-eat commercial menus (S.W. Foodtech., Co., Ltd.) such as hard-boiled whole eggs, scramble eggs, stewed eggs, omelets, etc., while the PS group received ready-to-eat commercial menus such as hard-boiled egg white or chicken sausages. On average, WE participants received 800 to 850 kcal/d, 2,100 to 2,260 mg of dietary cholesterol, 70 to 80 grams of protein, while PS participants received an additional 810 to 850 kcal/d, 50 to 220 mg of dietary cholesterol, and 70 to 80 grams of protein during the five school days. Participants in the control group received standard school lunches according to the Thai school lunch program. No group received additional meals or supplementation on weekends. Participants recruited in this study were followed for 35 weeks and seen on nine occasions by study research staff and dietitians.

### Outcomes

#### Anthropometric measurements

The main outcome of this long-term study was to determine the improvement of anthropometric indices. The anthropometries, biochemical indices and gut microbiomes of the participants were monitored at week 0 (baseline), week 14, and week 35 by trained researchers. Body weight was measured without shoes to the nearest 0·1 kg (Tanita HD-395, Tanita Corporation, Tokyo, Japan) and standing height was measured to the nearest 0·5 cm. software (Institute of Nutrition, Mahidol University, Thailand). Body mass index (BMI) was calculated for all participants. Weight and height data were converted to percentiles for weight for age (W/A), height for age (H/A) and weight for height (W/H) using Thai Growth program software (version 1·05, INMU, Nakhon Pathom, Thailand).^38^ Furthermore, subpopulations have also been characterized according to nutritional status, including underweight, stunting and wasting, which were defined as Z < −1.5 standard deviations (SD), to ensure that the intervention can promote weight and height, especially in the underweight and stunted groups. Stunting was defined as H/A Z < −1.5SD, underweight W/A Z < −1.5SD, and wasted W/H Z < −1.5SD.

#### Food record

At least 25% of the participants in each group were invited to participate in semi-structured face-to-face food recall and validated questionnaires with dietitians at their school for three times during the study period, including the first semester, end of the semester, and the second semester. To monitor compliance, the behavior and dietary intake of the children were obtained from a 3-day dietary record (two weekdays and one weekend).^39^ Participants provided the time, method of preparation, brand and weight of all the food ingredients and drinks consumed. The energy and nutrient intakes of all foods and drinks reported in each recall were summed to estimate the observed intakes of complementary feeding during the recall day. The nutrients, proteins, carbohydrates and fats were controlled by the Thai school lunch program (The National Electronics and Computer Technology Center (NECTEC), Pathumthani, Thailand) to ensure that all groups were adequate and there were no differences. Finally, energy intake, macronutrients, and micronutrients were calculated from the dietary records using INMUCAL–Nutrient Software version 4.0 (The Institute of Nutrition, Mahidol University (INMU), Nakhon Pathom, Thailand).

#### Specimen collection

Fasting blood samples were taken from participants for DNA extraction (**Supplement Methods 1 and 2**) and hematological and biochemical measurements at the start and end of the follow-up time points. Complete blood count (CBC), prealbumin, albumin, total cholesterol (TC), triglyceride (TG), high-density lipoprotein cholesterol (HDL-C) and calculated low-density lipoprotein cholesterol (LDL-C [based on Friedewald formula]^40^) were quantified in an accredited clinical laboratory (Siriraj Hospital, Bangkok, Thailand). Additionally, 15 grams of feces were randomly collected in 25% of the participants and placed in a 50 ml centrifuge tube at the beginning of the study (week 0) and the follow-up time points (week 14 and week 35), stored under cool conditions^41^, then aliquoted and frozen at −20°C for subsequent analyses.

Microbial DNA was isolated from 250 mg of feces using a QIAamp PowerFecal Pro DNA Kit (QIAGEN, Hilden, Germany) (**Supplement Method 3**). The samples were sent to the Centre d’expertise et de Services Génome Québec (Génome Québec, Montréal, Canada) for 16S rRNA sequencing. The gut microbiome study used the NovaSeq 6000 platform and Illumina sequencing by synthesis (SBS) to generate low error rate amplicon data. The analysis focused on V4 of the 16s rRNA genes, where 515F-806R was used as a primer. AmpliconSeq sequencing was performed on the NovaSeq platform (Génome Québec, Montréal, Canada). The raw sequence reads were processed using QIIME2 version 2021·4 and operated according to the standard pipeline recommended by Hall and Beiko.^42^ Briefly, the primers were trimmed and sequences with a quality score lower than 30 were filtered and further analyzed by merging the forward and reverse reads. The chimera was defined and removed by testing at least six abundances as potential parents. Taxonomy was classified with at least 97% sequence similarity to the Silva v132 database. The amplicon sequence variant (ASV) with an abundance of less than two sequences was filtered out. Phylogenetic analysis was performed by aligning the ASVs using MAFFT and a tree with FastTree. Finally, alpha and beta diversity and differential abundance were analyzed and visualized using the R package (details were described in Supplement Method 3). We generated a principal coordinate analysis (PCoA) using the weighted UniFrac distances between the first (T1) and the last time point (T3) of each group. The analysis of the composition of microbiomes with bias correction (ANCOM-BC) was applied to analyze the differential abundance in this study. We used pairwise comparisons with false discovery rate (FDR) correction to estimate the differences in taxonomic composition between the time points of each host group.

### Statistical analyses

Intention-to-treat analyses were applied for all inference analyses. Prespecified analyses were performed in three subgroups, as defined by characteristics at randomization: age, sex, W/A, H/A, and W/H. Continuous variables were expressed as mean ± standard deviation (SD) and discrete variables were expressed as percentages of the number of participants in each group. A chi-squared test was used to assess demographic characteristics and anthropometric data. For repeated measurements, the data were presented as mean ± SD and the *P*-value was obtained from the generalized estimation equation (GEE). Significant differences were defined as a *P*-value less than 0·05. Statistical analyses were performed using STATA version 17·0 (Stata Corporation, College Station, TX, USA).

#### Role of the funding source

The study sponsor had no role in study design, data collection, data analysis, data interpretation or writing the report. All authors had full access to all data in the study and the corresponding author had final responsibility for the decision to submit for publication.

## Results

This study was carried out from January 2019 to February 2020 (prior to COVID-19 pandemic). After recruitment, a total of 635 participants were randomly selected and allocated (**Figure 1** [238 for WE, 200 for PS, and 197 for Control]). The baseline characteristics and laboratory results were similar in the three groups. (**Table 1)**. The ratios between the number of boy and girl participants were similar at approximately 1: 1. At the baseline, the overall average age of participants was 9·8 ± 1·4 years old. Most of the participants had normal W/A, H/A, W/H. The mean percentiles of W/A, H/A, and W/H for all groups were not significantly different at *P*<0·05. Approximately 12-21% of the participants were underweight and 14-22% were stunted; in contrast, the proportion of overweight and obese participants was over 12%, 6%, and 70% had low prealbumin levels and low vitamin D levels, respectively. These results indicate that about a third of this population faced malnutrition of macronutrient or micronutrients. Based on Hb / Hct, the prevalence of anemia was estimated at 10% and more than two-thirds of them had a microcytic characteristic (MCV <80 fL). This could imply an iron deficiency state in this participant. Plasma lipid and lipoprotein concentrations, including TC, HDL-C, and LDL-C, were not significantly different between the groups, except for TG. The loss to follow-up was 46 participants (7%) due to illness, relocation, blood draw problems, or personal reasons (**Fig.1**).

**Table 1:** Baseline characteristics of participants.

| Variables | Control<br>[n=197] | PS<br>[n=200] | WE<br>[n=238] |
| --- | --- | --- | --- |
|  | n (%) | n (%) | n (%) |
| Age, mean ± SD, year | 9.2 ± 0.1 | 9.5 ± 0.1 | 9.6 ± 0.3 |
| Sex |  |  |  |
| Male | 103 (52.3) | 97 (48.5) | 108 (45.4) |
| Female | 94 (47.7) | 103 (51.5) | 130 (54.6) |
| Career of parents |  |  |  |
| Government officials | 5 (18.5) | 9 (33.3) | 13 (48.1) |
| Self-employment | 23 (26.1) | 27 (30.7) | 38 (43.2) |
| Agriculturist | 55 (34.6) | 45 (28.3) | 59 (37.1) |
| Company employee | 10 (16.7) | 22 (36.7) | 28 (46.7) |
| Unemployed | 9 (22.0) | 12 (29.3) | 20 (48.8) |
| Others (i.e., contractor) | 71 (39.9) | 52 (29.2) | 55 (30.9) |
| Weight, mean ± SD, kg | 31.6 ± 9.5 | 31.6 ± 8.1 | 32.1 ± 9.4 |
| Height, mean ± SD, cm | 137.1 ± 8.8 | 137.8 ± 9.3 | 138.7 ± 9.0 |
| W/A, mean ± SD, percentile | 103.7 ± 27.9 | 100.3 ± 22.8 | 103.4 ± 26.7 |
| Underweight | 24 (12.2) | 42 (21.0) | 40 (16.8) |
| Overweight | 19 (9.6) | 11 (5.5) | 12 (5.0) |
| H/A, mean ± SD, percentile | 100.1 ± 4.5 | 100.1 ± 4.4 | 100.3 ± 5.1 |
| Stunted | 29 (14.7) | 44 (22.0) | 41 (17.2) |
| W/H, mean ± SD, percentile | 102.3 ± 18.5 | 99.2 ± 14.7 | 102.2 ± 19.4 |
| Wasted | 24 (12.2) | 37 (18.5) | 21 (8.8) |
| Obese | 25 (12.7) | 36 (18.0) | 17 (7.1) |
| Blood pressure, mean ± SD, mm Hg | | | |
| Systolic | 103.2 ± 9.1 | 103.7 ± 9.1 | 104.2 ± 9.8 |
| Diastolic | 69.6 ± 5.6 | 70.7 ± 5.4 | 70.6 ± 5.9 |
| Hemoglobin, mean ± SD, g/dL | 12.8 ± 1.0 | 12.9 ± 1.0 | 12.9 ± 1.2 |
| <11.5 | 21 (10.7) | 19 (9.5) | 24 (10.1) |
| Hematocrit, mean ± SD, % | 39.2 ± 2.8 | 39.5 ± 1.8 | 39.7 ± 3.4 |
| < 35 | 17 (8.6) | 20 (10.0) | 24 (10.1) |
| MCV, mean ± SD, fL | 78.2 ± 5.6 | 78.5 ± 5.0 | 78.4 ± 6.3 |
| <80 | 125 (91.2) | 126 (63.0) | 152 (63.9) |
| Fasting glucose, mean ± SD, mg/dL | 86.6 ± 9.4 | 89.9 ± 8.3 | 86.8 ± 8.9 |
| Transferrin, mean ± SD, mg/dL | 257.2 ± 25.5 | 261.3 ± 30.3 | 262.0 ± 31.0 |
| Prealbumin, mean ± SD, mg/dL | 21.0 ± 3.2 | 21.2 ± 3.3 | 21.5 ± 3.2 |
| <16 | 12 (6.1) | 12 (6.0) | 13 (5.5) |
| Albumin, mean ± SD, g/dL | 4.4 ± 0.3 | 4.3 ± 0.3 | 4.4 ± 0.2 |
| Blood lipid level, mean ± SD, mg/dL | | | |
| TC | 178.2 ± 24.5 | 174.1 ± 28.1 | 175.9 ± 27.5 |
| TG | 76.8 ± 24.3 | 77.4 ± 26.1 | 79.5 ± 29.6 |
| HDL-C | 55.6 ± 11.0 | 55.1 ± 11.5 | 54.1 ± 9.9 |
| LDL-C | 105.1 ± 20.6 | 102.6 ± 22.8 | 104.5 ± 23.5 |
| Vitamin D, mean ± SD, ng/mL | 28.3 ± 6.3 | 24.9 ± 7.5 | 26.4 ± 6.6 |
| <30, % (95% CI) | 60.4 (57.1 – 63.2) | 75.0 (71.0 – 78.0) | 75.0 (72.0 – 77.0) |
| 20-29, % (95% CI) | 58.33 (54.1 – 61.2) | 53.19 (51.9 – 56.7) | 58.33 (53.9 – 60.1) |
| <20, % (95% CI) | 2.08 (1.01 – 3.34) | 16.67 (14.8 – 18.8) | 23.40 (20.9 – 25.5) |
| IGF-1, ng/mL, mean ± SD | 219.7 ± 111.1 | 265.7 ± 126.2 | 275.5 ± 137.6 |

Data are n (%), and mean ± SD. PS=protein substitute group. WE=whole egg group. W/A=weight for age. H/A=height for age. W/H=weight for height. MCV=mean corpuscular volume. TC=total cholesterol. TG=triglyceride. HDL-C=high-density lipoprotein cholesterol. LDL-C=low-density lipoprotein cholesterol. IGF-1=insulin-like growth factor 1.

### Consumption of whole eggs improved growth

At week 35, the results of child growth improved markedly and malnutrition, including undernutrition and overnutrition, improved in WE and PS compared to Control across almost all anthropometric measures **(Table 2** and **Supplement Table S1)**. We observed significant increases in weight and height in WE compared to PS and Control beginning at week 14 and noticeably at week 35. Participants in the WE group markedly gained a mean of 21·7 ± 13·5 % (4·4 ± 13·7 kg), while participants in the PS and control groups gained a mean of 20·9 ± 15·2 % (3·6 ± 13·5 kg) and 19·5 ± 12·4% (3·6 ± 13·3 kg), respectively (WE vs. PS; *P*<0.001, WE vs. control group; *P*<0·001). The height in WE increased by 24·6 ± 8·5 % (6·9 ± 13·8 cm), while the height in PS and Control increased by 22·7 ± 9·7 % (3·7 ± 13·6 cm), and 21·6 ± 9·3 % (3·4 ± 13·5 cm), respectively (WE vs. PS; *P*<0·001, WE vs. control group; *P*<0·001, [**Figure 2]**). Again, this increased growth in WE was significantly higher than the reference value recommended by the WHO for children of that age group. No significant differences in weight or height were observed between PS and Control after the intervention. In a subpopulation analysis **(Figure 3)**, a higher proportion of participants in WE than in PS and Control dramatically improved underweight, stunting, and wasting by 37-41%, 39-47%, and 35-44% (vs. PS [26-36%, 22-36%, and 27-31%] and Control [24-37%, 16-37%, and 26-38%]), respectively. Furthermore, children with overweight, tall stature, and obesity grew more in both the WE and PS groups than in the control group. WE had a greater improvement in H/A and W/A while PS had a remarkable improvement in weight but not in height (**Figure 3**).

**Table 2:**
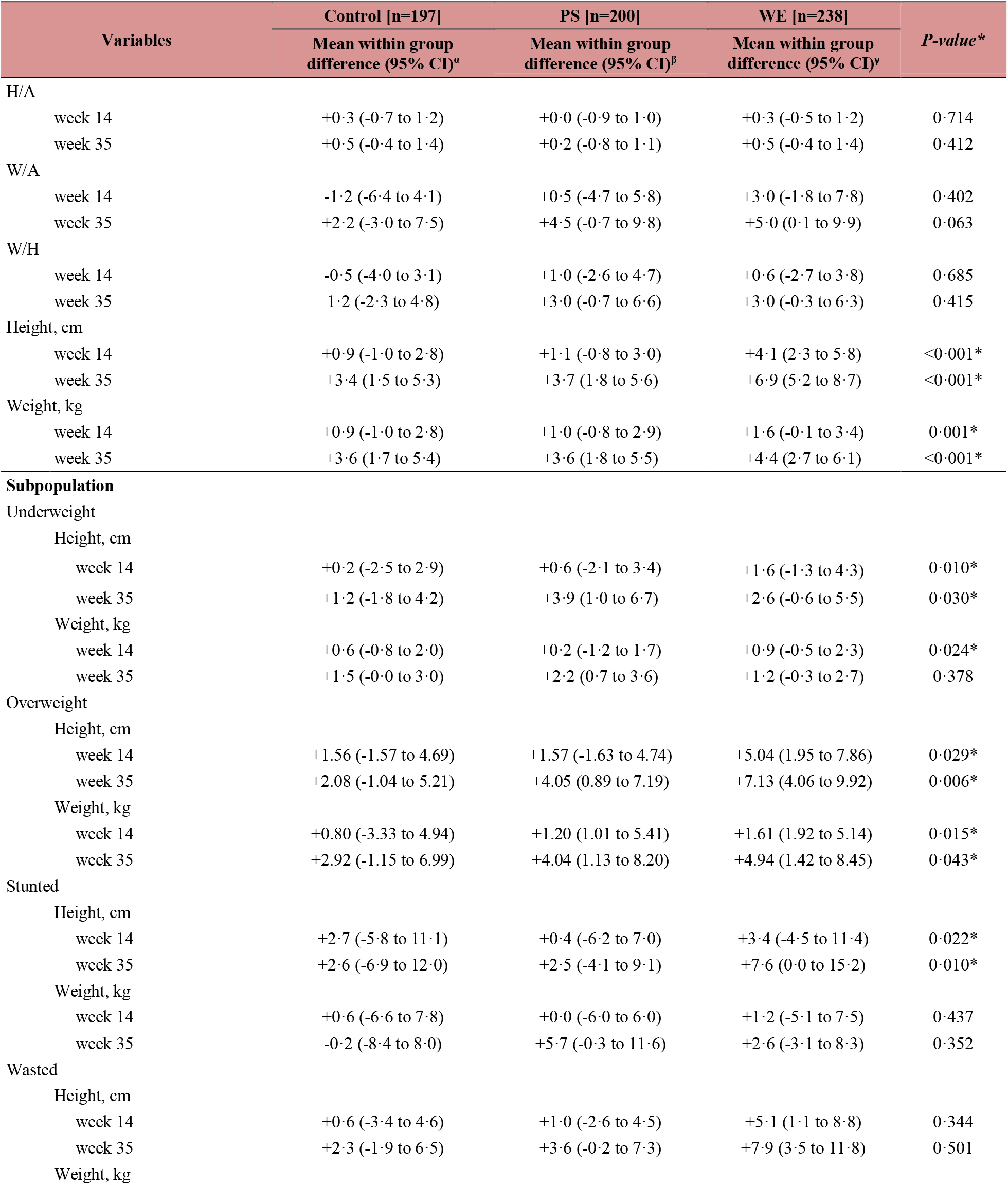
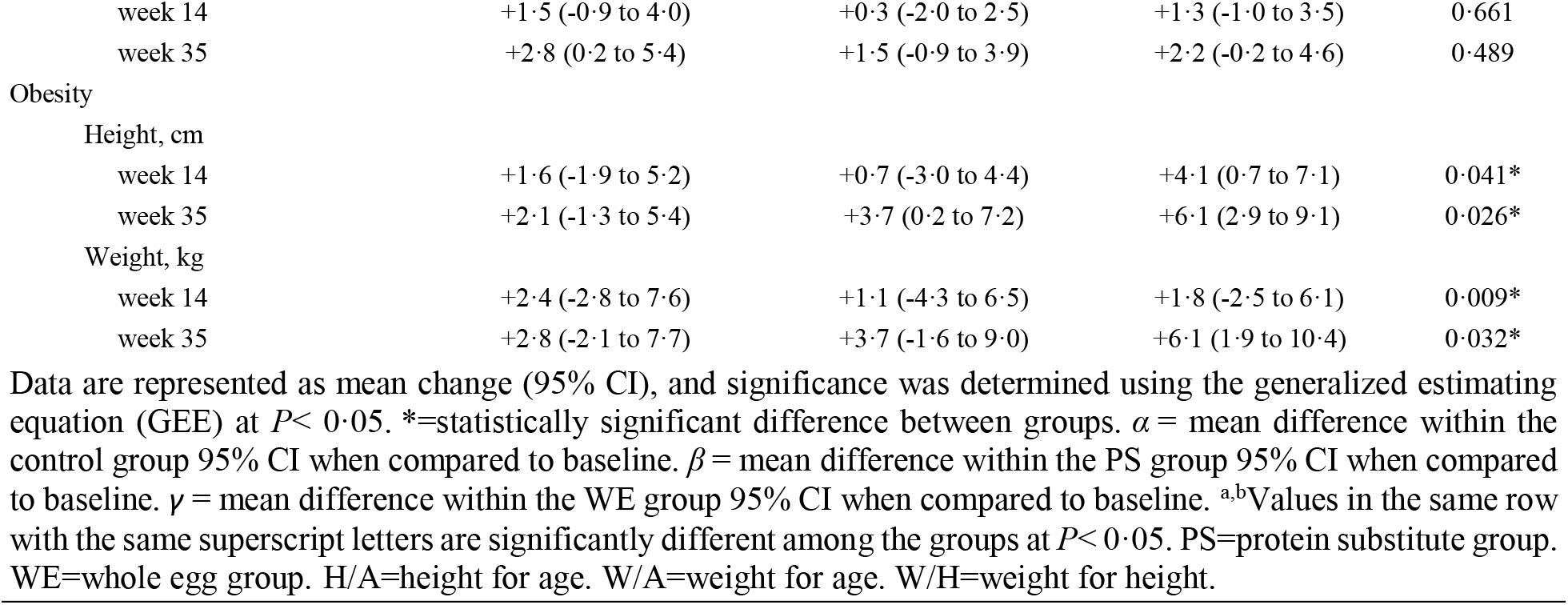
14 and 35 weeks change estimates for anthropometric of participants.

**Figure 2:**
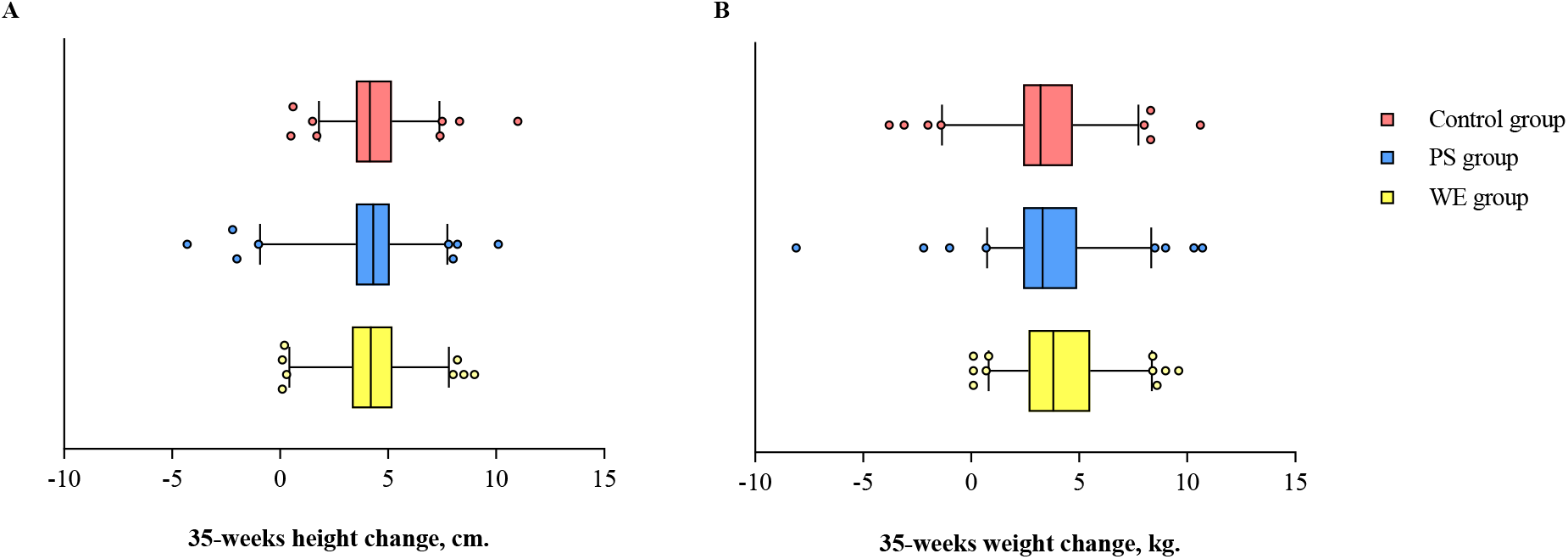
Body height and weight change in study group. (A) Mean of changes in height. (B) Mean of changes in weight. The bar graph represents the mean. Error bar indicate SD. PS=protein substitute group. WE=whole egg group.

**Figure 3:**
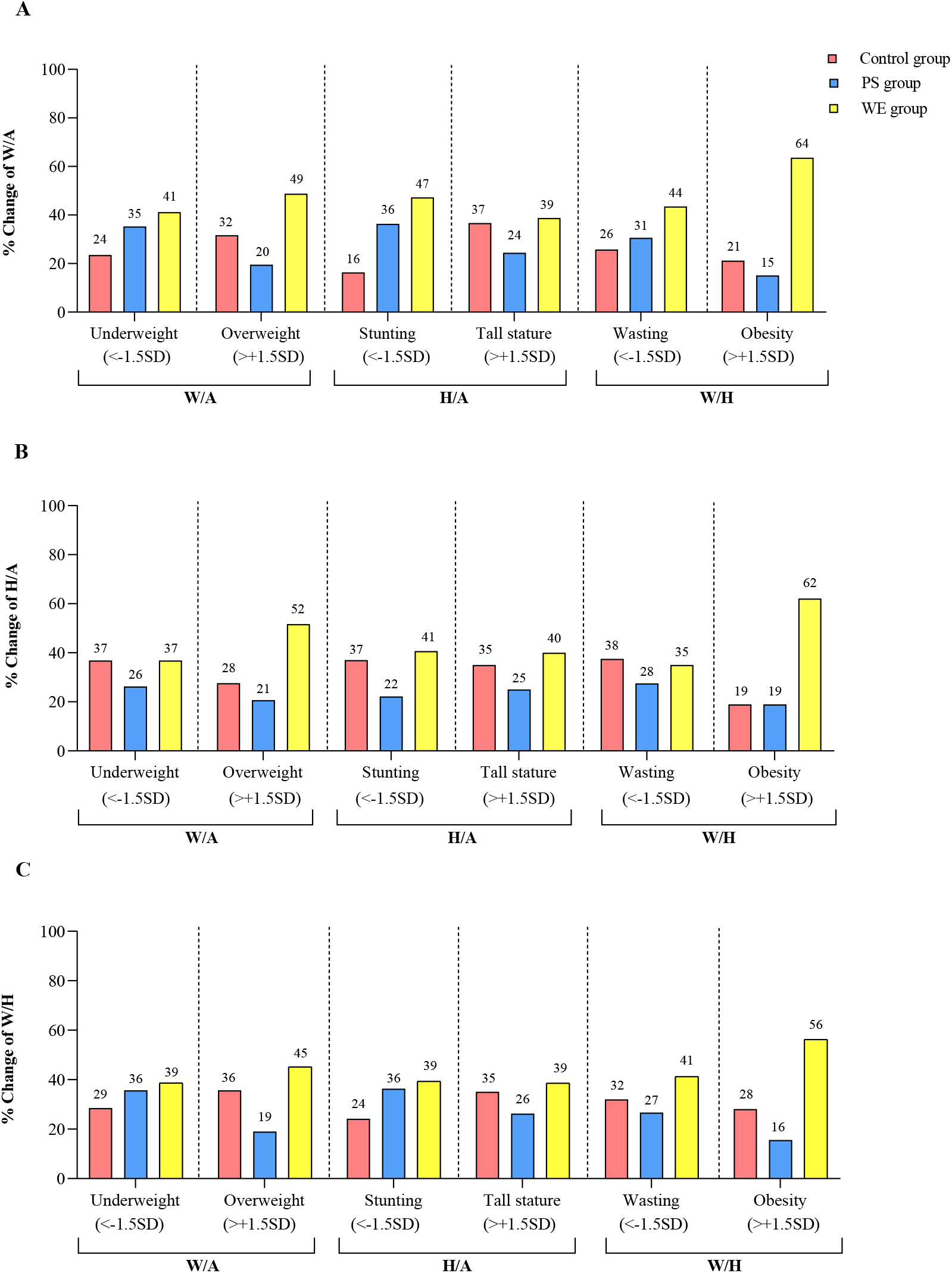
Body height and weight change in subpopulation. (A) Percentage of height change. (B) Percentage of weight change. The bar graph represents the percentage. PS=protein substitute group. WE=whole egg group.

### Plasma protein and anemia indices

At baseline, prealbumin levels <16 mg/dL, as a sensitive indicator of low nutritional status, were found in 5%, 6% and 6% in the WE, PS and control groups, respectively. Plasma concentrations of both prealbumin increased significantly by 1·3 mg/dL (95% CI, 0·7 to 1·9) in WE compared to the PS and control groups at week 14 and 35 (*P*<0·001 [**Table 3** and **Supplement Table S2**]). Iron deficiency and anemia are recognized as common nutritional deficits and signs of Thalassemia traits and other hemoglobinopathies in Thailand. Either hemoglobin less than 11·2 g/dL or hematocrit less than 35 was classified as anemia.^43^ We found an overall prevalence of anemia in 11% (70 participants), and 59% were boys and 41% were girls. Of 70 participants who were anemic, 57 (81%) had microcytic anemia (MCV <80 fL).

**Table 3:**
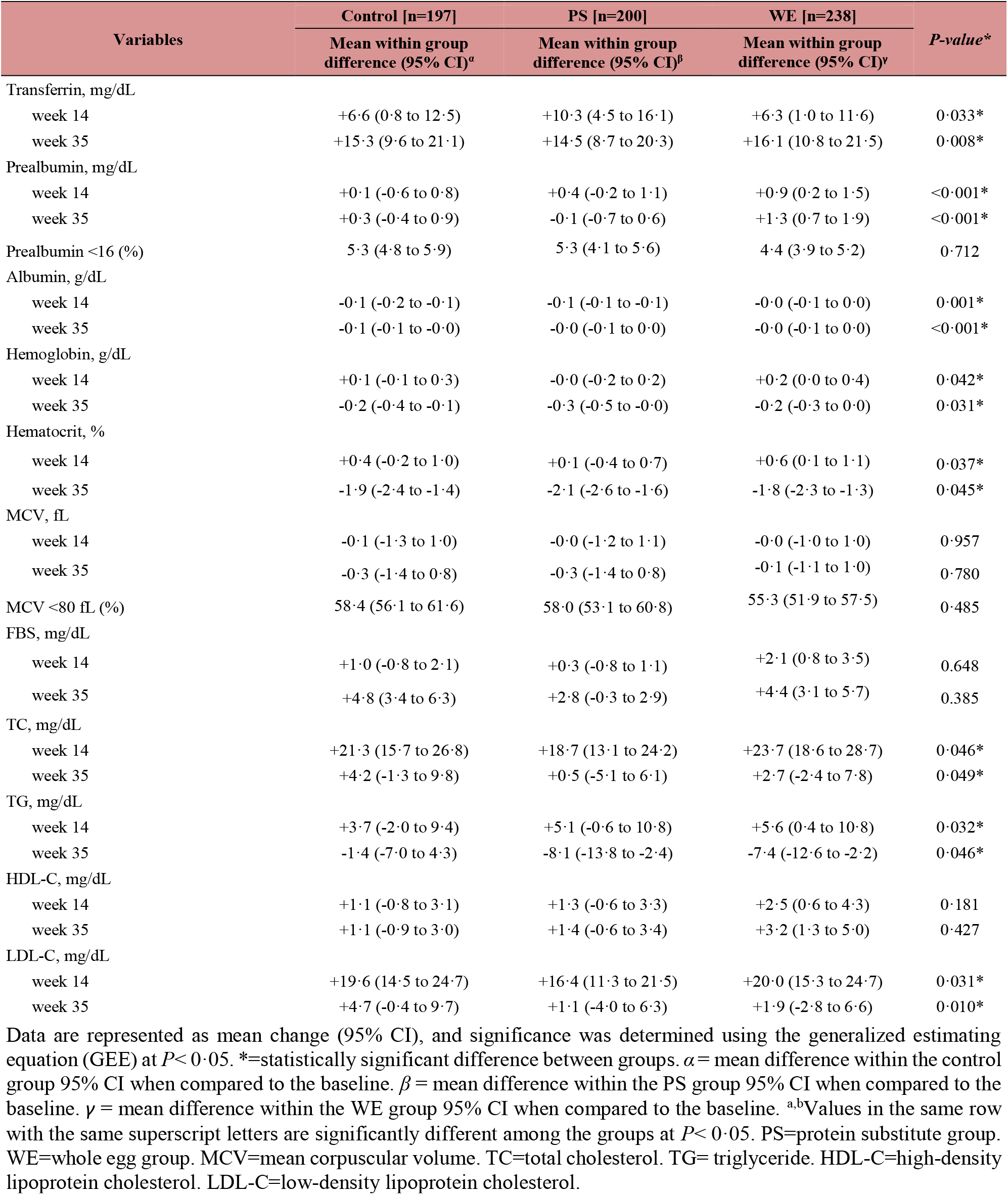
14 and 35 weeks change estimates for biochemical indices of participants.

| Variables | Control [n=197] | PS [n=200] | WE [n=238] | *P*-value* |
| --- | --- | --- | --- | --- |
|  | Mean within group difference (95% CI)<sup>α</sup> | Mean within group difference (95% CI)<sup>β</sup> | Mean within group difference (95% CI)<sup>γ</sup> |  |
| Transferrin, mg/dL |  |  |  |  |
| week 14 | +6.6 (0.8 to 12.5) | +10.3 (4.5 to 16.1) | +6.3 (1.0 to 11.6) | 0.033* |
| week 35 | +15.3 (9.6 to 21.1) | +14.5 (8.7 to 20.3) | +16.1 (10.8 to 21.5) | 0.008* |
| Prealbumin, mg/dL |  |  |  |  |
| week 14 | +0.1 (-0.6 to 0.8) | +0.4 (-0.2 to 1.1) | +0.9 (0.2 to 1.5) | <0.001* |
| week 35 | +0.3 (-0.4 to 0.9) | -0.1 (-0.7 to 0.6) | +1.3 (0.7 to 1.9) | <0.001* |
| Prealbumin <16 (%) | 5.3 (4.8 to 5.9) | 5.3 (4.1 to 5.6) | 4.4 (3.9 to 5.2) | 0.712 |
| Albumin, g/dL |  |  |  |  |
| week 14 | -0.1 (-0.2 to -0.1) | -0.1 (-0.1 to -0.1) | -0.0 (-0.1 to 0.0) | 0.001* |
| week 35 | -0.1 (-0.1 to -0.0) | -0.0 (-0.1 to 0.0) | -0.0 (-0.1 to 0.0) | <0.001* |
| Hemoglobin, g/dL |  |  |  |  |
| week 14 | +0.1 (-0.1 to 0.3) | -0.0 (-0.2 to 0.2) | +0.2 (0.0 to 0.4) | 0.042* |
| week 35 | -0.2 (-0.4 to -0.1) | -0.3 (-0.5 to -0.0) | -0.2 (-0.3 to 0.0) | 0.031* |
| Hematocrit, % |  |  |  |  |
| week 14 | +0.4 (-0.2 to 1.0) | +0.1 (-0.4 to 0.7) | +0.6 (0.1 to 1.1) | 0.037* |
| week 35 | -1.9 (-2.4 to -1.4) | -2.1 (-2.6 to -1.6) | -1.8 (-2.3 to -1.3) | 0.045* |
| MCV, fL |  |  |  |  |
| week 14 | -0.1 (-1.3 to 1.0) | -0.0 (-1.2 to 1.1) | -0.0 (-1.0 to 1.0) | 0.957 |
| week 35 | -0.3 (-1.4 to 0.8) | -0.3 (-1.4 to 0.8) | -0.1 (-1.1 to 1.0) | 0.780 |
| MCV <80 fL (%) | 58.4 (56.1 to 61.6) | 58.0 (53.1 to 60.8) | 55.3 (51.9 to 57.5) | 0.485 |
| FBS, mg/dL |  |  |  |  |
| week 14 | +1.0 (-0.8 to 2.1) | +0.3 (-0.8 to 1.1) | +2.1 (0.8 to 3.5) | 0.648 |
| week 35 | +4.8 (3.4 to 6.3) | +2.8 (-0.3 to 2.9) | +4.4 (3.1 to 5.7) | 0.385 |
| TC, mg/dL |  |  |  |  |
| week 14 | +21.3 (15.7 to 26.8) | +18.7 (13.1 to 24.2) | +23.7 (18.6 to 28.7) | 0.046* |
| week 35 | +4.2 (-1.3 to 9.8) | +0.5 (-5.1 to 6.1) | +2.7 (-2.4 to 7.8) | 0.049* |
| TG, mg/dL |  |  |  |  |
| week 14 | +3.7 (-2.0 to 9.4) | +5.1 (-0.6 to 10.8) | +5.6 (0.4 to 10.8) | 0.032* |
| week 35 | -1.4 (-7.0 to 4.3) | -8.1 (-13.8 to -2.4) | -7.4 (-12.6 to -2.2) | 0.046* |
| HDL-C, mg/dL |  |  |  |  |
| week 14 | +1.1 (-0.8 to 3.1) | +1.3 (-0.6 to 3.3) | +2.5 (0.6 to 4.3) | 0.181 |
| week 35 | +1.1 (-0.9 to 3.0) | +1.4 (-0.6 to 3.4) | +3.2 (1.3 to 5.0) | 0.427 |
| LDL-C, mg/dL |  |  |  |  |
| week 14 | +19.6 (14.5 to 24.7) | +16.4 (11.3 to 21.5) | +20.0 (15.3 to 24.7) | 0.031* |
| week 35 | +4.7 (-0.4 to 9.7) | +1.1 (-4.0 to 6.3) | +1.9 (-2.8 to 6.6) | 0.010* |

Data are represented as mean change (95% CI), and significance was determined using the generalized estimating equation (GEE) at $P < 0.05$ . \* = statistically significant difference between groups. $\alpha$ = mean difference within the control group 95% CI when compared to the baseline. $\beta$ = mean difference within the PS group 95% CI when compared to the baseline. $\gamma$ = mean difference within the WE group 95% CI when compared to the baseline. <sup>a,b</sup>Values in the same row with the same superscript letters are significantly different among the groups at $P < 0.05$ . PS = protein substitute group. WE = whole egg group. MCV = mean corpuscular volume. TC = total cholesterol. TG = triglyceride. HDL-C = high-density lipoprotein cholesterol. LDL-C = low-density lipoprotein cholesterol.

### Cardiometabolic Variables

TC, TG, HDL levels markedly increased at week 14 compared to baseline in all groups (*P*<0· 05), while HDL levels increased significantly only in the WE group but not in the PS and control groups at week 14. Subsequently, at week 35, TC levels returned to similar levels in all groups compared to baseline (ns), while TG levels showed a marked decrease in the PS and WE groups, but not in the control group, compared to baseline and week 14 (*P*<0·05). Surprisingly, HDL levels increased in the WE group at week 35 (3·2 mg/dL (95%CI 1·3 to 5·0 [*P* = 0·001]). No significant differences in LDL-C concentration were observed in all groups. However, the mean HDL-C concentration at week 35 did not show significant differences in WE group compared to the PS and control groups, but had trend increases in the WE group (57·3 ± 8·0 mg/dL) as compared to PS (56·5 ± 10·2 mg/dL) and the control group (56·7 ± 10·0 mg/dL) (WE vs. PS; *P* = 0·410, WE vs. C; *P* = 0·510) shown in **Table 3, Figure 4**, and **Supplement Table S2**.

**Figure 4:**
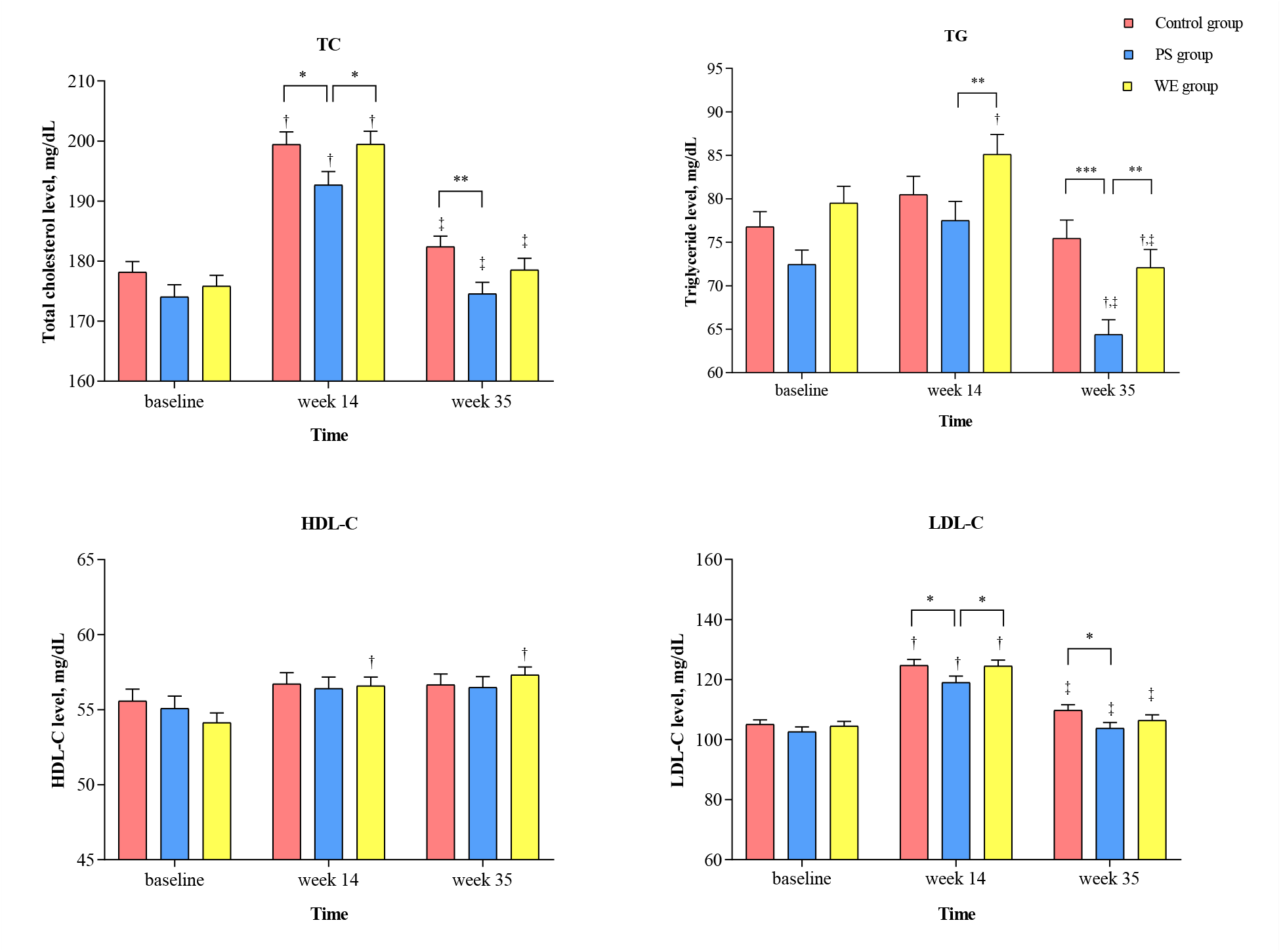
Plasma lipid levels change in study group. (A) TC level, (B) TG level (C) LDL-C level and (D) HDL-C level. The bar graph represents the mean. Error bar indicate SEM. ^*^ The statistical significance between group at *P*<0·05 ^**^ The statistical significance between group at *P*<0·01 ^***^ The statistical significance between group at *P*<0·001 ^†^ The statistical significance within group when compared to baseline ^‡^ The statistical significance within group when compared to week 14 PS=protein substitute group. WE=whole egg group. TC=total cholesterol. TG=triglyceride. LDL-C=low density lipoprotein cholesterol. HDL-C=high density lipoprotein cholesterol.

### Overall energy intake

No significant differences were observed in the overall mean dietary energy intake and macronutrients, including carbohydrates, protein, fat, and fiber, except cholesterol, between the groups during the study period **(Table 4)**. Significant differences in cholesterol levels (mg / day) were observed in WE (368·5 ± 92·4 mg/day) as compared to the PS (230·3 ± 62·6 mg/day) and the control group (236·9 ± 65·2 mg/day), (*P*<0·001). The comparison of nutrition intake between the groups on weekdays and weekends showed that the average energy intake was not significantly different between the groups on weekdays, while it was significantly different on weekends. For macronutrients (i.e., carbohydrates, protein and fat), especially cholesterol, there was a significant difference between the groups on both weekdays and weekends (**Supplement Table 3**).

**Table 4:** Average nutrient intake of participants during the study period.

| Energy/Nutrients | control [n=39] | PS [n=41] | WE [n=45] | *P-value* |
| --- | --- | --- | --- | --- |
| | Mean ± SD | Mean ± SD | Mean ± SD | |
| Energy (kcal/day) | 1029.2 ± 143.5 | 1021.8 ± 129.3 | 1018.0 ± 126.5 | 0.343 |
| Carbohydrate (g/day) | 124.1 ± 19.8 | 117.9 ± 17.9 | 116.0 ± 17.0 | 0.345 |
| Protein (g/day) | 41.7 ± 7.4 | 46.6 ± 6.7 | 44.8 ± 7.0 | 0.136 |
| Fat (g/day) | 40.7 ± 7.8 | 40.4 ± 6.7 | 41.7 ± 7.0 | 0.486 |
| Saturated fatty acids (g/day) | 11.6 ± 2.9 | 11.8 ± 2.8 | 11.1 ± 2.8 | 0.607 |
| Cholesterol (mg/day) | 236.9 ± 65.2<sup>a</sup> | 230.3 ± 62.6<sup>a</sup> | 368.5 ± 92.4<sup>b</sup> | <0.001* |
| Dietary fiber (g/day) | 4.2 ± 1.9 | 3.7 ± 1.0 | 3.8 ± 5.4 | 0.880 |
| Sodium (mg/day) | 1559.4 ± 378.9 | 1512.6 ± 326.6 | 1456.1 ± 308.8 | 0.656 |
| Calcium (mg/day) | 386.8 ± 111.4 | 366.4 ± 89.8 | 377.9 ± 113.7 | 0.923 |
| Iron (mg/day) | 5.1 ± 1.1 | 5.4 ± 1.0 | 5.6 ± 1.3 | 0.098 |
| Zinc (mg/day) | 3.0 ± 0.7 | 3.4 ± 0.6 | 3.2 ± 0.6 | 0.174 |
| Vitamin A, RAE (μg) | 274.0 ± 143.0 | 281.0 ± 165.4 | 446.0 ± 291.3 | 0.042* |
| Beta-carotene (mcg/day) | 395.6 ± 246.5 | 383.2 ± 228.9 | 409.4 ± 240.2 | 0.506 |
| Thiamin (mg/day) | 0.8 ± 0.3 | 1.0 ± 1.1 | 0.8 ± 0.3 | 0.980 |
| Riboflavin (mg/day) | 0.8 ± 0.2 | 1.0 ± 0.2 | 0.9 ± 0.2 | 0.435 |
| Niacin (mg/day) | 7.1 ± 1.7 | 7.9 ± 1.8 | 7.2 ± 1.7 | 0.111 |
| Vitamin C (mg/day) | 22.6 ± 13.2 | 19.1 ± 12.0 | 16.5 ± 9.3 | 0.776 |

Data are represented as mean ± SD, and significance was determined using repeated measures ANOVA at $P < 0.05$ .
<sup>a,b</sup>Values in the same row with the same superscript letters are significantly different among the groups at $P < 0.05$ .
PS=protein substitute group. WE=whole egg group. RAE= retinol activity equivalents.

However, our preliminary data showed that average energy intake, most macronutrients and micronutrients, and vitamins were significantly higher in all groups on weekdays than on weekends. The WE group had a higher frequency of carbohydrate and protein intake (including vegetables and other animal products) intake than PS and control groups. The number of snacks and desserts was lower in WE than in the PS and control groups.

### Taxonomic classification at the beginning of the study and at the end of the intervention between the groups

A total of 455 658 ASVs were detected, corresponding to 2 kingdoms, 29 phyla, 61 classes, 137 orders, 233 families, and 519 genera. Of the nine genera with the highest abundance in the host group (**Figure 5**), there was a significant change in relative abundance between baseline and week 35 in WE. The *Bifidobacterium* that was found to have a positive effect on child growth in undernourished children^44^ increased up to 1·28-fold and *Prevotella* increased 2·63-fold and 2·68-fold in the WE group and in the control groups, respectively. We age-matched children with poor growth to children with normal growth to obese ones and compared the change in abundance of ASVs over time. After egg supplementation in WE, *Prevotella* increased, as reported in an earlier study.^45^ The ASVs classified by taxonomy from the previous step were imported into the R platform. We used the observed ASVs, Chao1 richness, Shannon index, and the InvSimpson index for the alpha diversity estimator. These were used to characterize the diversity of species among groups. PCoA was generated from weighted UniFrac distances for the beta diversity analysis. Bacterial diversity in the WE, PS, and control groups did not significantly change alpha or beta diversity **(Figure 6)**. Taxonomically classified ASVs were differentially abundant by ANCOM-BC using the R package. **Figure 7** shows the logarithmic ratio of abundance at the last time/ first time point. The genera with higher abundances after supplementation represent a positive direction in the bar graph. On the contrary, genera with lower abundance after supplementation were represented in a negative direction. The abundance of *Agathobacter, Candidatus Soleaferrea*, and *Clostridia* vadinBB60 was significantly increased in the control group. *Enterobacteriaceae* decreased significantly in the control group. Furthermore, the abundance of *genera of Eubacterium Ventriosum, Anaerofilum*, and *Incertae Sedis* increased significantly in the control and PS groups.

**Figure 5:**
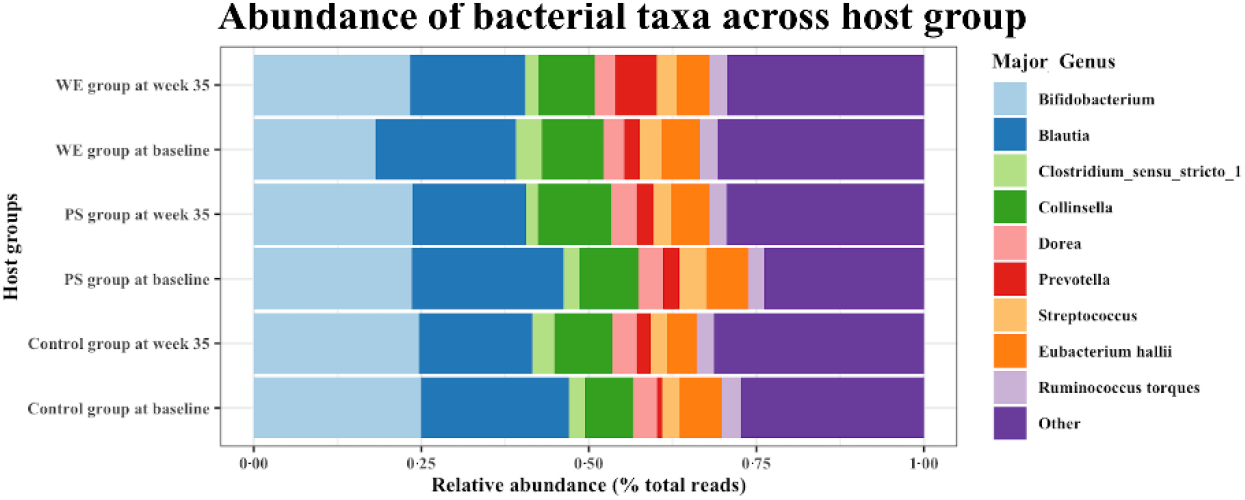
Taxonomy classification in study group. Stacked column graph represents the relative abundance of majority bacterial genera across the host group. PS=protein substitute group. WE=whole egg group.

**Figure 6:**
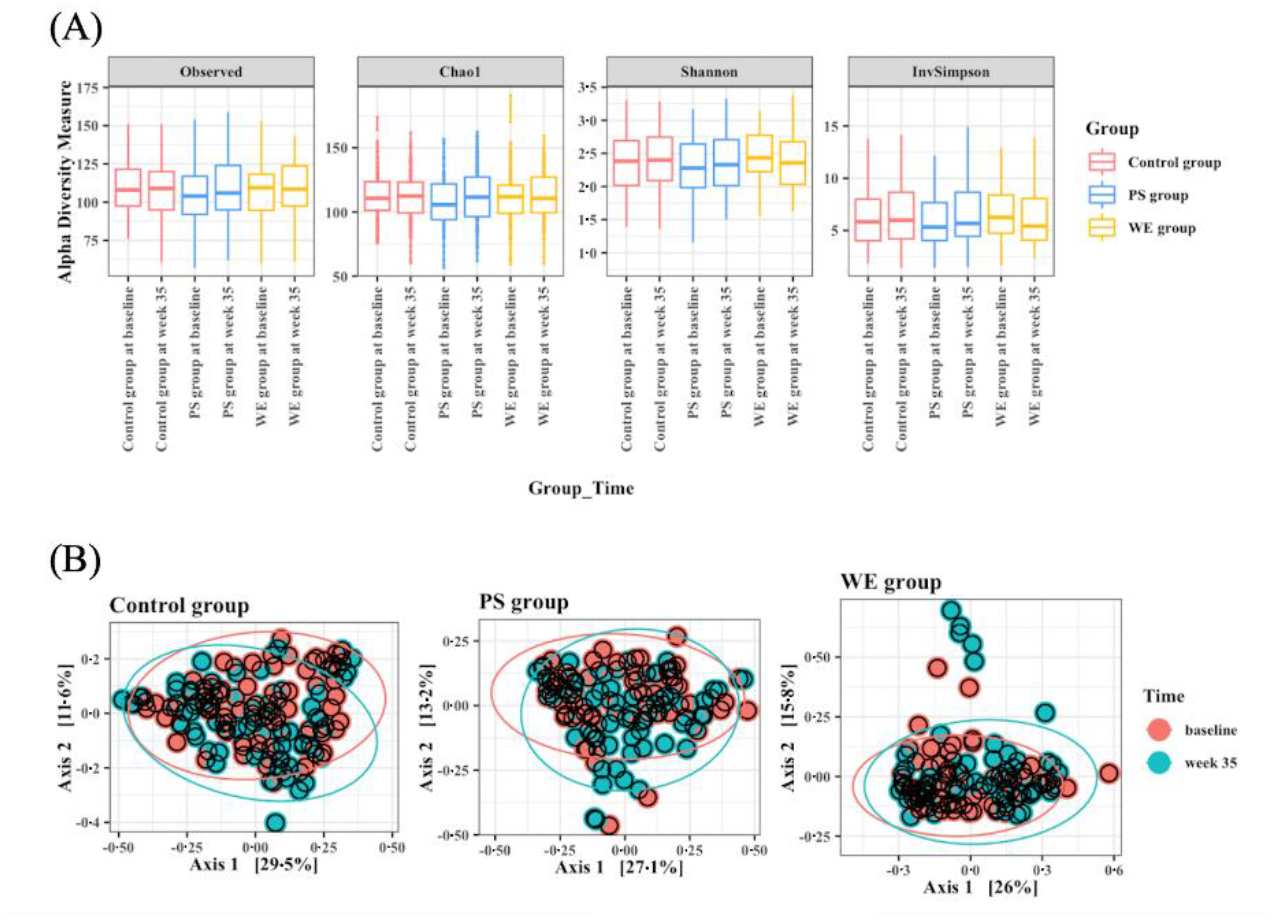
Bacterial diversity. (A) Box-plots of alpha diversity observed in different time points among the host group. (B) Multidimensional scaling plot of beta diversity described by Permutational multivariate analysis of variance (PERMANOVA) and the Bray-Curtis dissimilarity measure. The color of the plot represents the time point. C=control group. PS=protein substitute group. WE=whole egg group. T1=baseline. T3=week 35.

**Figure 7:**
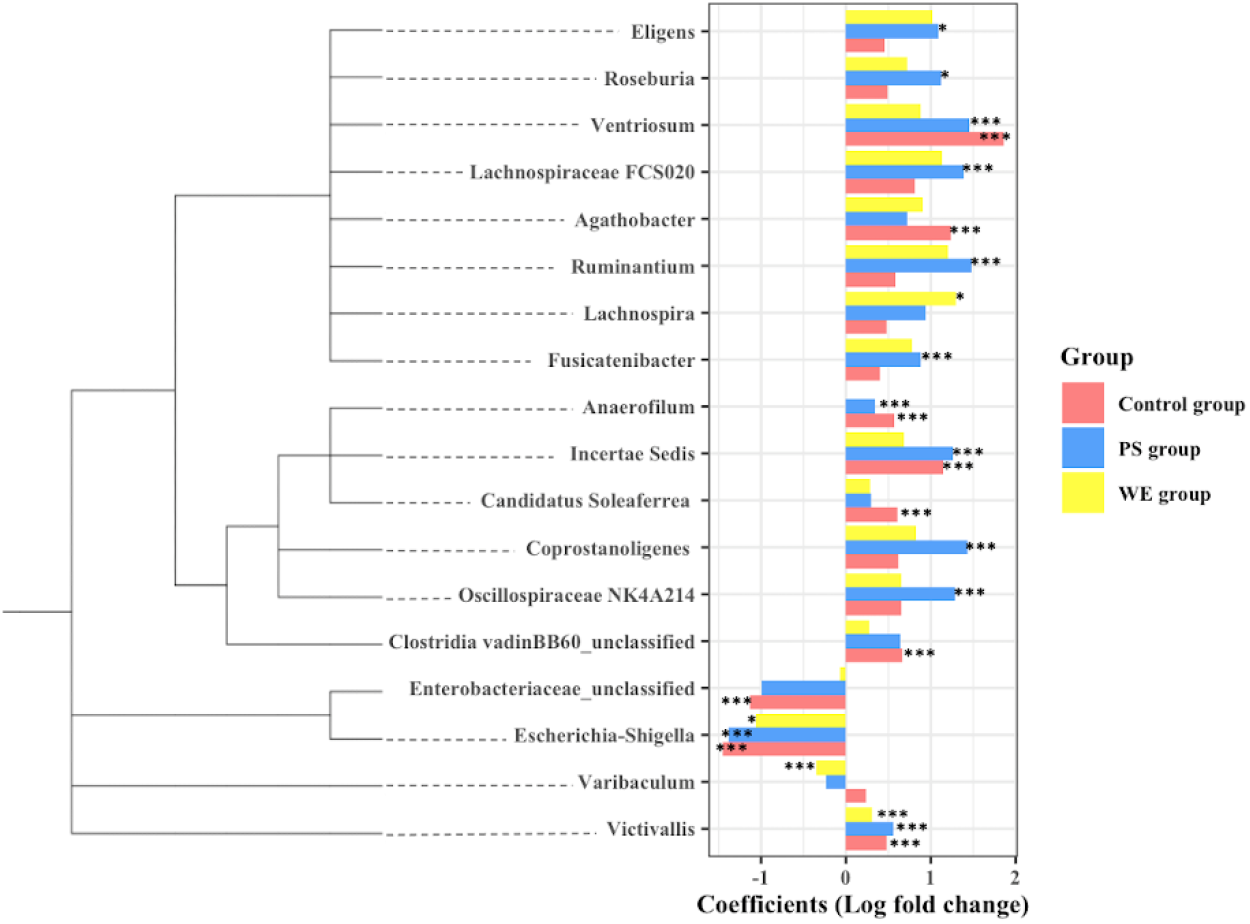
Bacterial differential abundance. The phylogenetics tree and differential analysis result of the genera with a significant difference after egg supplemented in each host group. Data are represented by effect size (log fold change of T3/T1) and 95% confidence interval bars (two-sided; FDR adjusted) derived from the ANCOM-BC model. All effect sizes with adjusted *P*< 0·05 are indicated, \**P*< 0·05, \*\**P*< 0·01, *** *P*< 0·001 of significance. C=control group. PS=protein substitute group. WE=whole egg group.

## Discussion

This RCT was the first long-term intervention that provided two additional whole eggs per school day for 35 weeks, beginning in the first semester and continuing through the second semester in multiple regions of Thailand, compared to the PS and control groups or a usual school lunch program. This was an extension of a 3-month egg intervention project described previously.^21^ We confirmed that this produced a significant positively biological impact upon adolescent growth, particularly improving stunting and underweight. This intervention was associated with improved biomarkers, including lipoproteins, microbiota and healthy dietary patterns in children.

The World Health Organization (WHO) and the United Nations Children’s Fund (UNICEF) reported the prevalence of stunting (height for age Z < −2SD from the median of WHO child growth standards) among children under 5 years of age (%) and a trend in child malnutrition that is greater than 10 to 50% in Africa, the eastern Mediterranean and South East Asia, including Thailand.^46^ We observed that more than 10% of rural primary school children were underweight, stunted or wasted, had low vitamin D levels, low prealbumin levels, or were anemic. These conditions involved both inadequate macronutrient and micronutrient intake. However, we did not observe different caloric intakes across study groups. Our results showed that additional egg consumption may influence healthier dietary patterns. In Thailand eggs are often eaten with rice, a filling meal that can reduce the need for snacks and desserts. In fact, a previous study in U.S. children showed that egg consumption was significantly associated with higher amounts of several nutrients, including protein, total and saturated fat, alpha-linolenic acid, DHA, lutein + zeaxanthin, choline, potassium, phosphorus, selenium, riboflavin, and vitamin D, A, and vitamin E.^47^ Similarly, a cross-sectional survey in the U.S. reported that eggs and foods containing eggs can be an important part of a healthy dietary pattern when balanced with other food rich in nutrients.^48^ Benjamin (2005) reported that more than 60% of households in middle-income countries had an underweight family member, with the majority being in developing countries undergoing a nutritional transition, and some countries were affected by a double burden of malnutrition.^49,50^ At present, the world is facing with socioeconomic inequality, which can lead to starvation and malnutrition. And the COVID-19 pandemic has even disproportionately affected economically disadvantaged groups. Although many low-cost commercial foods are high in calories; in contrast, they often have poor nutrient profiles so called “poor nutrition-dense foods”.

This study is important for the health policy of primary-school children. This finding is consistent and confirms a randomized controlled trial by Iannotti et al. that reported that egg consumption significantly improved growth in young children.^8^ In Ecuador, one egg per day for six months was reported to have reduced stunting by 47% and increased linear growth by 0·63 length-for-age Z (LAZ).^51^ In a cohort of rural children in western Kenya, Mosites et al. showed that the height gain of the child was associated with milk and egg consumption. This finding aligns with evidence that eggs provide proteins and micronutrients that can promote child growth.^52^ Furthermore, we found that whole egg supplementation increased protein in the blood and decreased concentrations of lipid profiles, including triglycerides, while HDL-C showed an increasing trend in egg supplementation. This is consistent with our previous study demonstrating that continuous egg consumption resulted in increased blood protein levels and high-density lipoprotein cholesterol (HDL-C), while low-density lipoprotein cholesterol (LDL-C) decreased. Similarly, daily egg consumption promotes HDL lipid composition and function.^53^ Likewise, egg consumption increases total cholesterol, LDL-C and HDL-C, but not LDL-C:HDL-C, TC:HDL-C, and TG, when compared to low egg control diets.^54^ Furthermore, Fernandez et al. reported that eating whole eggs increases the size of HDL lipoprotein particles and increases the activity of lecithin-cholesterol acyltransferase (LCAT).^22,23^ Recently, US cohort studies and meta-analysis data showed that moderate egg consumption (up to one egg per day) is not associated with a potentially lower risk of cardiovascular disease in Asian populations.^55^ However, the association between egg intake and CVD risk remains unclear, which may be associated with the size of lipoprotein particles and the activity of the enzyme in lipid metabolism.^23,56^

We evaluated the changes in the gut microbiome structure after whole egg supplementation. We observed increased levels of *Bifidobacterium* in the group supplemented with whole eggs. *Bifidobacterium* is a human milk oligosaccharide (HMO) used by bacteria.^57^ They are considered to have health-promoting benefits in humans, especially in the infant gut.^58^ These microbes produce a variety of metabolites, including lactic, acetic, propionic, and butyric acids, which benefit the host’s immune system.^57^ On the contrary, a decrease in this microbiota has been associated with a high incidence of diseases, such as irritable bowel syndrome (IBS)^59^ and autoimmune hepatitis.^60^ In Thai children, abundance is negatively correlated with the consumption of fish, beef, and bread.^61^ However, the relationship between these genera and the consumption of vitamins and minerals remains unclear. In our study, the abundance of *Lachnospira* was significantly higher after whole-egg supplementation. *Lachnospira* are anaerobic, fermentative and chemoorganotrophic.^62^ Normally, this genus is well known as one of the SCFA producers throughout the whole grain fermenter.^63^ Vanegas et al. supplemented 81 healthy adults with whole or refined grains for six weeks and the abundance of *Lachnospira* increased significantly. However, there were no significant changes in the inflammatory responses or microbial products.^64^ Our results showed that the abundance of *Varibaculum* was significantly lower after whole-egg supplementation. Furthermore, there is little evidence of the relationship between *Varibaculum* and host health at the genus level. Kang et al. reported that the abundance of *Varibaculum* was significantly higher in patients with invasive cervical cancer (CAN) compared to healthy controls.^65^

This research has strengths that suggest that its findings may have important implications for public policy. First, this is a large-scale, one-year randomized controlled trial. We collected rural schoolchildren, including central, eastern, and western Thailand, homogenized by geographical and food patterns. We randomly assigned participants to study groups according to their weight and age to ensure that they did not have significantly different nutritional statuses, such as being underweight or overweight. Second, we used tools for the evaluation of food intake, including the 3-day record and the 24-hour recall. Additionally, we designed food records over three time periods to achieve a high level of precision of nutrition data. Third, this study showed an important verified discovery that tackling malnutrition, especially in low-middle-income communities, could be achievable by using locally available high-quality proteins such as eggs, milk and chicken. This affects clinically, biologically and physiologically related microbiota as well as impacts upon healthier food choices and children’s behaviors. However, we included only rural school areas about two to three hours’ drive from Bangkok and so our results may not be representative of the entire child population of Thailand.

We conclude that long-term whole egg supplementation significantly increases growth and improves important biomarkers in young children without adverse effects on blood cholesterol levels. Our results suggest that whole egg supplementation also promotes intestinal microbial diversity by maintaining intestinal microbiota composition that benefits health. More information is needed on the mechanistic effects of egg consumption on gut microbiota and growth. Whole egg supplementation is a feasible, low-cost, and effective intervention to address the problem of protein-energy malnutrition in school-age children.

## Data Availability

All data produced in the present work are contained in the manuscript.

## Contributions

SS, AS, and KM conceived and designed the study, analyzed the data, and drafted the manuscript. PP, BP, and KS revised and reviewed the nutrition data. PP, SK, SS, and IT reviewed and analyzed the microbiome data. A.S. performed the biostatistical analyses and reviewed the data. The SPHERE group (SS, PP, BP, AS, TM, SO, TP, and SP) performed patient care and revised and reviewed the manuscript accordingly. The corresponding author (KM) attests that all listed authors meet the authorship criteria and that no others that meet the criteria have been omitted. All authors have read the manuscript and agreed with its content.

## Declaration if interests

The authors declare that they have no conflict of interest.

## Acknowledgments

The authors thank all parents and children for their participation. We thank all of the teachers in each school for helping to manage egg consumption among the subjects. We thank the members of SPHERE staff, T. A. Central Lab staff, Siriraj Applied Thai Traditional Medicine staff, and research assistants for their recruitment and data collection. The authors are also grateful to the management of the Siriraj Medical Research Center for providing the office space and biobanks. We thank S.W. Foodtech., Co., Ltd. for providing the eggs used in this study.

## Notes

### Competing Interest Statement

The authors have declared no competing interest.

### Clinical Trial

NCT04896996

### Clinical Protocols

https://clinicaltrials.gov/ct2/show/NCT04896996

### Author Declarations

Institutional Review Board of Siriraj Hospital, Mahidol University (COA No. Si 322/2017)

