## Supplement for "Effects of Long-Term Supplementation of Eggs on Growth, Biochemical Indices, and Microbiota of Rural Thai Primary School Children"

### Supplementary Appendix

This appendix has been provided by the authors to provide readers with additional information about this work.

**Contents:**


| Contents: | Page |
|---|---|
| Method S1: Plasma and serum collection | 2 |
| Method S2: Human DNA extracted from blood sample | 2 |
| Method S3: Bacterial DNA extracted from stool sample | 2 |
| Table S1: Mean change estimates for anthropometric of participants | 4 |
| Table S2: Mean change estimates for biochemical indices of participants | 6 |
| Table S3: Comparison of nutrition intake between groups by day of subjects | 8 |
| Table S4. Comparison of nutrition intake within groups by day of subjects | 10 |
| Table S5. Food pattern of subjects | 11 |
| Figure S1: Food pattern in the population studied | 12 |

**Method S1: Plasma and serum collection**

1. Venous blood (6 mL) into an EDTA tube (treated with an anticoagulant) for plasma and into a clotted blood tube for serum.
2. For serum, put sample tube at room temperature for 30 minutes before centrifugation.
3. All plasma and serum were centrifuged at 3,000 rpm at 4 ° C for 10 minutes by centrifugator.
4. Aliquot plasma and serum
5. Storage at -20 ° C (analysis within 1 month) and -80 ° C for study in the future.

**Method S2: Human DNA extracted from blood sample**

An Automated Chemagic 360 instrument (CMG-1074, PerkinElmer, Baesweiler, Germany) was used. Briefly, 2 mL of whole blood was collected into a deep well, and 15 µL of protein kinase was added to each well. Preparation of reagents and instruments according to the manufacturer's instructions. The DNA concentration and a purity ratio (acceptable in range 1·8-1·9) were measured using FLUOstar Omega (software version 5·5 R4, BMG LABTECH, Ortenberg, Germany).

**Method S3: Bacterial DNA extracted from stool sample**

The QIAamp® PowerFecal® Pro DNA Kit (QIAGEN, Hilden, Germany) was used (following to the manufacturer). In a PowerBead Pro Tube, 250 mg of stool was weighed, and 800 µL of solution CD1. Vortex using PowerLyzer 24 Homogenizer at maximum speed for 10 min. and centrifuged at 15000 g for 1 min. Pipette 600 µl. of the supernatant into a microcentrifuge tube and 200 µL of solution CD2 was added after centrifugation. Transfer 700 µL of supernatant to a microcentrifuge tube and add 600 µL of solution CD3. The lysate (650 µL) was loaded into the MB spin column and centrifuged. The flow-through was discarded, the MB spin column was placed into a collection tube, and 500 µl of EA solution was added. The flow-through was then centrifuged and discarded. Then, 500 µL of solution C5 was added to the MB spin column, centrifuged, and the flow-through discarded. The MB spin column was carefully centrifuged for 2 min at 16 000 g. Finally, 50 µL of solution C6 and centrifuge at 15 000 × g for 1 min. The MB spin column was discarded. DNA is currently ready for downstream applications. Microbial DNA concentration and a purity ratio (acceptable in range 1·8-1·9) were measured using FLUOstar Omega (software version 5·5 R4, BMG LABTECH, Ortenberg, Germany).

The samples were sent to the Centre d'expertise et de Services Génome Québec (Génome Québec, Montréal, Canada) for 16S rRNA sequencing. The gut microbiome study used the NovaSeq 6000 platform and Illumina sequencing by synthesis (SBS). Phylogenetic analysis was performed by aligning the ASVs using MAFFT and a tree with FastTree. Further, alpha and beta diversity and differential abundance were analyzed and visualized using the R package. Alpha diversity describes the bacterial variety within each sample, and ASV richness describes the number of genera in each subject (richness). The Chao1 richness estimates the number of unobserved genera. The Shannon index describes the richness and relative abundance (evenness). The InvSimpson index is the inverse of the classic Simpson diversity estimator and describes the probability that two randomly selected sequences belong to the same genus. Beta diversity describes the divergence between individual hosts.

| Variables | Control [n=197] | PS [n=200] | WE [n=238] | *P-value* |
|---|---|---|---|---|
| | **Mean ± SD** | **Mean ± SD** | **Mean ± SD** | |
| H/A | | | | |
| baseline | 100·1 ± 4·5 | 100·1 ± 4·4 | 100·3 ± 5·1 | 0·399 |
| week 14 | 100·4 ± 4·6 | 100·1 ± 4·4 | 100·6 ± 5·3 | 0·714 |
| week 35 | 100·6 ± 2·5 | 100·3 ± 4·7 | 100·8 ± 5·1 | 0·412 |
| change | +0·5 | +0·2 | +0·5 | |
| W/A | | | | |
| baseline | 103·7 ± 27·9 | 100·3 ± 22·8 | 103·4 ± 26·7 | 0·317 |
| week 14 | 102·6 ± 26·0 | 100·9 ± 24·5 | 106·4 ± 29·2 | 0·402 |
| week 35 | 106·0 ± 27·1 | 104·9 ± 27·3 | 108·4 ± 30·2 | 0·063 |
| change | +2·2 | +4·5 | +5·0 | |
| W/H | | | | |
| baseline | 102·3 ± 18·5 | 99·2 ± 14·7 | 102·2 ± 19·4 | 0·688 |
| week 14 | 101·9 ± 18·0 | 100·3 ± 16·6 | 102·7 ± 19·8 | 0·685 |
| week 35 | 103·5 ± 18·1 | 102·2 ± 18·0 | 105·1 ± 20·7 | 0·415 |
| change | +1·2 | +3·0 | +3·0 | |
| Height, cm | | | | |
| baseline | 137·1 ± 8·8 | 137·8 ± 9·3 | 138·7 ± 9·0 | 0·358 |
| week 14 | 138·0 ± 9·7[a] | 138·9 ± 9·9[a] | 142·7 ± 10·0[b] | <0·001* |
| week 35 | 140·5 ± 9·5[a] | 141·5 ± 9·9[a] | 145·6 ± 10·3[b] | <0·001* |
| change | +3·4 | +3·7 | +6·9 | |
| Weight, kg | | | | |
| baseline | 31·6 ± 9·5[a] | 31·6 ± 8·1[a] | 34·3 ± 9·3[b] | 0·016* |
| week 14 | 32·5 ± 9·5[a] | 32·6 ± 8·8[a] | 35·9 ± 9·9[b] | 0·001* |
| week 35 | 35·2 ± 10·4[a] | 35·2 ± 9·6[a] | 38·7 ± 11·0[b] | <0·001* |
| change | +3·6 | +3·6 | +4·4 | |
| **Subpopulation** | | | | |
| Underweight | | | | |
| Height, cm | | | | |
| baseline | 131·8 ± 13·5 | 133·4 ± 14·5 | 134·4 ± 13·6 | 0·246 |
| week 14 | 132·0 ± 13·5[a] | 134·1 ± 15·3[ab] | 136·0 ± 15·1[b] | 0·010* |
| week 35 | 133·0 ± 15·6[a] | 137·3 ± 17·8[b] | 137·0 ± 14·8[b] | 0·030* |
| change | +1·2 | +3·9 | +2·6 | |
| Weight, kg | | | | |
| baseline | 24·7 ± 6·7[a] | 25·4 ± 7·2[ab] | 26·3 ± 7·7[b] | 0·018* |
| week 14 | 25·3 ± 6·3[a] | 25·6 ± 7·8[a] | 27·2 ± 8·2[b] | 0·024* |
| week 35 | 26·2 ± 8·0 | 27·6 ± 9·1 | 27·5 ± 9·2 | 0·378 |
| change | +1·5 | +2·2 | +1·2 | |
| Overweight | | | | |
| Height, cm | | | | |
| baseline | 142·8 ± 14·9 | 143·0 ± 18·9 | 143·5 ± 15·0 | 0·757 |
| week 14 | 144·3 ± 14·3[a] | 144·6 ± 18·4[a] | 148·5 ± 14·9[b] | 0·029* |
| week 35 | 144·8 ± 12·3[a] | 147·1 ± 18·5[a] | 150·6 ± 14·4[b] | 0·006* |
| change | +2·1 | +4·1 | +7·1 | |
| Weight, kg | | | | |
| baseline | 46·0 ± 23·2[ab] | 42·8 ± 19·8[a] | 47·5 ± 19·0[b] | 0·020* |
| week 14 | 46·8 ± 22·9[ab] | 44·0 ± 19·4[a] | 49·1 ± 18·7[b] | 0·015* |
| week 35 | 48·9 ± 23·3[ab] | 46·8 ± 19·5[a] | 52·4 ± 20·0[b] | 0·043* |
| change | +2·9 | +4·0 | +4·9 | |
| Stunted | | | | |
| Height, cm | | | | |

| | | | | |
|---|---|---|---|---|
| baseline | 120·0 ± 35·3 | 125·0 ± 46·7 | 127·3 ± 50·7 | 0·277 |
| week 14 | 122·7 ± 32·8[a] | 125·4 ± 45·3[ab] | 130·8 ± 48·5[b] | 0·022* |
| week 35 | 122·6 ± 20·4[a] | 127·5 ± 43·5[a] | 135·0 ± 36·4[b] | 0·010* |
| change | +2·6 | +2·5 | +7·6 | |
| Weight, kg | | | | |
| baseline | 21·8 ± 17·5 | 22·2 ± 22·5 | 25·8 ± 29·8 | 0·445 |
| week 14 | 22·3 ± 26·1 | 22·2 ± 24·3 | 27·0 ± 39·2 | 0·437 |
| week 35 | 21·6 ± 17·7 | 27·7 ± 67·9 | 28·4 ± 33·3 | 0·352 |
| change | -0·2 | +5·7 | +2·6 | |
| Wasted | | | | |
| Height, cm | | | | |
| baseline | 138·8 ± 19·5 | 137·6 ± 18·7 | 136·4 ± 17·8 | 0·419 |
| week 14 | 139·5 ± 21·7 | 138·7 ± 19·3 | 141·5 ± 20·5 | 0·344 |
| week 35 | 141·2 ± 22·4 | 141·2 ± 20·8 | 144·3 ± 23·1 | 0·501 |
| change | +2·3 | +3·6 | +7·9 | |
| Weight, kg | | | | |
| baseline | 26·4 ± 12·7 | 27·1 ± 11·4 | 27·9 ± 12·6 | 0·392 |
| week 14 | 27·9 ± 13·1 | 27·4 ± 11·6 | 29·2 ± 12·6 | 0·661 |
| week 35 | 29·2 ± 13·5 | 28·7 ± 11·8 | 30·1 ± 14·6 | 0·489 |
| change | +2·8 | +1·5 | +2·2 | |
| Obesity | | | | |
| Height, cm | | | | |
| baseline | 141·6 ± 16·3 | 139·4 ± 20·8 | 141·0 ± 15·6 | 0·436 |
| week 14 | 143·2 ± 16·9[ab] | 140·1 ± 20·8[a] | 145·0 ± 17·0[b] | 0·041* |
| week 35 | 143·7 ± 15·3[a] | 143·1 ± 19·0[ab] | 147·1 ± 17·3[b] | 0·026* |
| change | +2·1 | +3·7 | +6·1 | |
| Weight, kg | | | | |
| baseline | 47·5 ± 27·0 | 43·1 ± 26·6 | 47·5 ± 22·6 | 0·386 |
| week 14 | 49·9 ± 29·0[a] | 44·2 ± 25·7[b] | 49·3 ± 23·8[a] | 0·009* |
| week 35 | 50·3 ± 26·5[ab] | 46·8 ± 24·2[b] | 53·6 ± 25·4[b] | 0·032* |
| change | +2·8 | +3·7 | +6·1 | |

Data are represented as mean ± SD, and significance was determined using the generalized estimating equation (GEE) at $P< 0·05$. *=statistically significant difference between group. $\alpha$ = mean difference within the control group 95% CI when compared to the baseline. $\beta$ = mean difference within the PS group 95% CI when compared to the baseline. $\gamma$ = mean difference within the WE group 95% CI when compared to the baseline. [a,b]Values in the same row with the same superscript letters are significantly different among the groups at $P<0·05$. PS=protein substitute with low cholesterol group. WE=whole egg group. H/A=height for age. W/A=weight for age. W/H=weight for height.

**Table S1: Mean change estimates for anthropometric of participants**

| Variables | Control [n=197] Mean ± SD | PS [n=200] Mean ± SD | WE [n=238] Mean ± SD | *P-value* |
|---|---|---|---|---|
| Transferrin, mg/dL | | | | |
| baseline | 257·2 ± 25·5 | 261·3 ± 30·3 | 262·3 ± 31·0 | 0·142 |
| week 14 | 263·9 ± 26·4[a] | 271·6 ± 30·0[b] | 268·6 ± 29·0[ab] | 0·033* |
| week 35 | 272·9 ± 30·0[a] | 275·8 ± 32·0[ab] | 278·5 ± 32·7[b] | 0·008* |
| change | +15·3 | +14·5 | +16·1 | |
| Prealbumin, mg/dL | | | | |
| baseline | 21·0 ± 3·2 | 21·2 ± 3·3 | 21·5 ± 3·2 | 0·771 |
| week 14 | 21·1 ± 3·2[a] | 21·7 ± 3·1[a] | 22·4 ± 3·7[b] | <0·001* |
| week 35 | 21·3 ± 3·4[a] | 21·2 ± 3·4[a] | 22·8 ± 4·5[b] | <0·001* |
| change | +0·3 | -0·1 | +1·3 | |
| Albumin, g/dL | | | | |
| baseline | 4·4 ± 0·3 | 4·4 ± 0·3 | 4·4 ± 0·2 | 0·420 |
| week 14 | 4·3 ± 0·1[a] | 4·3 ± 0·1[a] | 4·4 ± 0·2[b] | 0·001* |
| week 35 | 4·3 ± 0·3[a] | 4·3 ± 0·3[a] | 4·4 ± 0·3[b] | <0·001* |
| change | -0·1 | -0·0 | -0·0 | |
| Hemoglobin, g/dL | | | | |
| baseline | 12·8 ± 1·0 | 12·9 ± 1·0 | 12·9 ± 1·2 | 0·757 |
| week 14 | 12·9 ± 0·8[ab] | 12·8 ± 0·9[a] | 13·1 ± 1·1[b] | 0·042* |
| week 35 | 12·5 ± 0·8[a] | 12·6 ± 0·9[a] | 12·8 ± 0·9[b] | 0·031* |
| change | -0·2 | -0·3 | -0·2 | |
| Hematocrit, % | | | | |
| baseline | 39·2 ± 2·8[a] | 39·5 ± 1·8[ab] | 39·7 ± 3·4[b] | 0·049* |
| week 14 | 39·6 ± 2·1[a] | 39·6 ± 2·6[a] | 40·3 ± 3·2[b] | 0·037* |
| week 35 | 37·3 ± 2·4[a] | 37·4 ± 2·4[a] | 37·9 ± 2·6[b] | 0·045* |
| change | -1·9 | -2·1 | -1·8 | |
| MCV, fL | | | | |
| baseline | 78·2 ± 5·6 | 78·5 ± 5·0 | 78·4 ± 6·3 | 0·755 |
| week 14 | 78·0 ± 5·8 | 78·5 ± 5·2 | 78·4 ± 6·5 | 0·957 |
| week 35 | 77·9 ± 5·5 | 78·2 ± 5·1 | 78·3 ± 6·5 | 0·780 |
| change | -0·3 | -0·3 | -0·1 | |
| FBS, mg/dL | | | | |
| baseline | 86·6 ± 9·4 | 87·6 ± 8·4 | 86·8 ± 9·0 | 0.234 |
| week 14 | 87·3 ± 7·2 | 87·9 ± 7·6 | 88·9 ± 10·3 | 0.648 |
| week 35 | 91·4 ± 7·8 | 90·4 ± 7·5 | 91·2 ± 9·1 | 0.385 |
| change | +4·8 | +2·8 | +4·4 | |
| TC, mg/dL | | | | |
| baseline | 178·2 ± 24·5 | 174·1 ± 28·1 | 175·9 ± 27·5 | 0·131 |
| week 14 | 199·4 ± 29·5[a] | 192·7 ± 31·4[b] | 199·5 ± 33·2[a] | 0·046* |
| week 35 | 182·4 ± 24·4[a] | 174·6 ± 26·9[b] | 178·6 ± 39·2[ab] | 0·049* |
| change | +4·2 | +0·5 | +2·7 | |
| TG, mg/dL | | | | |
| baseline | 76·8 ± 24·3[ab] | 72·5 ± 23·5[a] | 79·5 ± 29·6[b] | 0·017* |
| week 14 | 80·5 ± 29·5[ab] | 77·5 ± 16·8[a] | 85·1 ± 35·5[b] | 0·032* |
| week 35 | 75·5 ± 29·6[a] | 64·4 ± 24·0[b] | 72·1 ± 31·8[a] | 0·046* |
| change | -1·4 | -8·1 | -7·4 | |
| HDL-C, mg/dL | | | | |
| baseline | 55·6 ± 11·0 | 55·1 ± 11·5 | 54·1 ± 9·9 | 0·604 |
| week 14 | 56·7 ± 10·4 | 56·4 ± 10·9 | 56·6 ± 9·0 | 0·181 |
| week 35 | 56·7 ± 10·0 | 56·5 ± 10·2 | 57·3 ± 8·0 | 0·427 |
| change | +1·1 | +1·4 | +3·2 | |

| | | | | |
|---|---|---|---|---|
| LDL-C, mg/dL | | | | |
| baseline | 105·1 ± 20·6 | 102·6 ± 22·8 | 104·5 ± 23·5 | 0·113 |
| week 14 | 124·8 ± 27·5[a] | 119·1 ± 29·7[b] | 124·5 ± 30·6[a] | 0·031* |
| week 35 | 109·8 ± 26·3[a] | 103·8 ± 27·0[b] | 106·5 ± 27·6[ab] | 0·010* |
| change | +4·6 | +1·2 | +1·9 | |

Data are represented as mean ± SD, and significance was determined using the generalized estimating equation (GEE) at $P$<0·05. *=statistically significant difference between group. $\alpha$ = mean difference within the control group 95% CI when compared to the baseline. $\beta$ = mean difference within the PS group 95% CI when compared to the baseline. $\gamma$ = mean difference within the WE group 95% CI when compared to the baseline. [a,b]Values in the same row with the same superscript letters are significantly different among the groups at $P$<0·05. PS=protein substitute with low cholesterol group. WE=whole egg group. MCV=mean corpuscular volume. FBS=fasting blood sugar. TC=total cholesterol. TG= triglyceride. HDL-C=high-density lipoprotein cholesterol. LDL-C=low-density lipoprotein cholesterol.

**Table S2: Mean change estimates for biochemical indices of participants**

| Energy/Nutrients | Control [n=39] | PS [n=41] | WE [n=45] | *P-value* |
|---|---|---|---|---|
| | Mean ± SD | Mean ± SD | Mean ± SD | |
| Energy (kcal/day) | | | | |
| weekday | 1067·4 ± 148·5 | 1034·9 ± 151·1 | 1044·2 ± 149·6 | 0·588 |
| weekend | 924·9 ± 154·7[a] | 1036·29 ± 157·3[b] | 958·2 ± 155·8[a] | 0·003 |
| Carbohydrate (g/day) | | | | |
| weekday | 129·0 ± 20·7[a] | 118·0 ± 21·0[b] | 113·8 ± 20·8[b] | 0·002 |
| weekend | 111·4 ± 20·6[a] | 124·8 ± 20·9[b] | 117·7 ± 20·7[ab] | 0·013 |
| Protein (g/day) | | | | |
| weekday | 42·8 ± 7·7[a] | 48·1 ± 7·8[b] | 47·2 ± 7·7[b] | 0·004 |
| weekend | 38·3 ± 8·6[a] | 44·4 ± 8·7[b] | 40·2 ± 8·7[a] | 0·004 |
| Fat (g/day) | | | | |
| weekday | 42·2 ± 7·9 | 41·2 ± 8·1 | 44·5 ± 8·0 | 0·131 |
| weekend | 36·2 ± 8·5[a] | 40·0 ± 8·6[b] | 36·3 ± 8·6[a] | 0·050 |
| Saturated fatty acid (g/day) | | | | |
| weekday | 12·4 ± 3·3 | 12·3 ± 3·3 | 11·8 ± 3·2 | 0·607 |
| weekend | 9·9 ± 3·2[ab] | 11·0 ± 3·3[a] | 9·6 ± 3·2[b] | 0·007 |
| Cholesterol (mg/day) | | | | |
| weekday | 240·3 ± 87·2[a] | 225·3 ± 88·8[a] | 430·0 ± 87·9[b] | <0·001* |
| weekend | 232·6 ± 73·8 | 242·3 ± 75·0 | 241·6 ± 74·4 | 0·800 |
| Dietary fiber (g/day) | | | | |
| weekday | 4·6 ± 1·8[a] | 3·8 ± 1·8[b] | 3·8 ± 1·7[b] | 0·050 |
| weekend | 3·4 ± 1·3 | 3·6 ± 1·2 | 3·6 ± 1·3 | 0·829 |
| Sodium (mg/day) | | | | |
| weekday | 1549·6 ± 385·1 | 1562·1 ± 391·6 | 1434·8 ± 387·9 | 0·224 |
| weekend | 1568·4 ± 388·9 | 1421·1 ± 395·5 | 1524·4 ± 391·8 | 0·207 |
| Calcium (mg/day) | | | | |
| weekday | 450·4 ± 121·7 | 409·1 ± 123·8 | 439·6 ± 122·6 | 0·273 |
| weekend | 236·4 ± 103·6[a] | 299·7 ± 105·4[b] | 244·9 ± 104·4[a] | 0·010 |
| Iron (mg/day) | | | | |
| weekday | 5·2 ± 1·3[a] | 5·7 ± 1·3[ab] | 5·9 ± 1·3[b] | 0·046 |
| weekend | 4·7 ± 1·3 | 5·1 ± 1·3 | 4·7 ± 1·3 | 0·219 |
| Zinc (mg/day) | | | | |
| weekday | 3·1 ± 0·1[a] | 3·5 ± 0·6[b] | 3·5 ± 0·7[b] | 0·001 |
| weekend | 2·8 ± 0·8[ab] | 3·1 ± 0·8[a] | 2·6 ± 0·9[b] | 0·013 |
| Vitamin A, RAE (µg) | | | | |
| weekday | 326·0 ± 693·3 | 386·6 ± 705·1 | 576·5 ± 698·4 | 0·205 |
| weekend | 207·3 ± 106·7[ab] | 238·7 ± 108·5[a] | 174·0 ± 107·5[b] | 0·017 |
| Beta-carotene (mcg/day) | | | | |
| weekday | 449·9 ± 287·4 | 376·9 ± 292·2 | 434·1 ± 289·5 | 0·473 |
| weekend | 276·9 ± 241·3[a] | 377·7 ± 245·4[ab] | 379·6 ± 243·0[b] | 0·007 |
| Thiamin (mg/day) | | | | |
| weekday | 0·7 ± 0·5 | 0·9 ± 0·5 | 0·7 ± 0·5 | 0·179 |
| weekend | 0·8 ± 1·1[ab] | 1·2 ± 1·1[a] | 0·8 ± 1·1[b] | 0·050 |
| Riboflavin (mg/day) | | | | |
| weekday | 0·9 ± 0·2[a] | 1·1 ± 0·2[b] | 1·0 ± 0·2[b] | <0·001* |
| weekend | 0·7 ± 0·3[a] | 0·9 ± 0·3[b] | 0·7 ± 0·2[a] | <0·001* |
| Niacin (mg/day) | | | | |
| weekday | 7·0 ± 1·9 | 7·8 ± 2·0 | 7·2 ± 2·0 | 0·157 |
| weekend | 7·0 ± 2·1[a] | 8·8± 2·2[b] | 7·0 ± 2·2[a] | <0·001* |
| Vitamin C (mg/day) | | | | |
| weekday | 24·3 ± 13·1[a] | 19·2 ± 13·3[ab] | 16·0 ± 13·2[b] | 0·012 |

| | | | | |
|---|---|---|---|---|
| weekend | 20·3 ± 13·5 | 19·1 ± 13·7 | 17·3 ± 13·6 | 0·581 |

Data are represented as mean ± SD, and significance was determined using repeated measures ANOVA at $P < 0·05$. [a,b]Values in the same row with the same superscript letters are significantly different among the groups at $P < 0·05$. PS=protein substitute with low cholesterol group. WE=whole egg group. RAE= retinol activity equivalents.

**Table S3: Comparison of nutritional intake between groups by day of participants**

| Energy/Nutrients | Control [n=39] | | *P-value* | PS [n=41] | | *P-value* | WE [n=45] | | *P-value* |
|---|---|---|---|---|---|---|---|---|---|
| | weekday | weekend | | weekday | weekend | | weekday | weekend | |
| Energy (kcal/day) | 1067·4 ± 148·5 | 924·9 ± 154·7 | <0·001* | 1034·9 ± 151·1 | 1036·2 ± 157·3 | 0·961 | 1044·2 ± 149·6 | 958·2 ± 155·8 | 0·001 |
| Carbohydrate (g/day) | 129·0 ± 20·7 | 111·4 ± 20·6 | <0·001* | 118·0 ± 21·0 | 124·8 ± 20·9 | 0·059 | 113·8 ± 20·8 | 117·7 ± 20·7 | 0·253 |
| Protein (g/day) | 42·8 ± 7·7 | 38·3 ± 8·6 | 0·004 | 48·1 ± 7·8 | 44·4 ± 8·7 | 0·016 | 47·2 ± 7·7 | 40·2 ± 8·7 | <0·001* |
| Fat (g/day) | 42·2 ± 7·9 | 36·2 ± 8·5 | <0·001* | 41·2 ± 8·1 | 40·0 ± 8·6 | 0·453 | 44·5 ± 8·0 | 36·3 ± 8·5 | <0·001* |
| Saturated fatty acid (g/day) | 12·4 ± 3·3 | 9·9 ± 3·2 | <0·001* | 12·3 ± 3·3 | 11·0 ± 3·3 | 0·049 | 11·8 ± 3·2 | 9·6 ± 3·2 | 0·001 |
| Cholesterol (mg/day) | 240·3 ± 87·2 | 232·6 ± 73·8 | 0·651 | 225·3 ± 88·8 | 242·3 ± 75·0 | 0·312 | 430·0 ± 87·9 | 241·6 ± 74·3 | <0·001* |
| Dietary fiber (g/day) | 4·6 ± 1·8 | 3·4 ± 1·3 | <0·001* | 3·8 ± 1·8 | 3·6 ± 1·2 | 0·441 | 3·8 ± 1·7 | 3·6 ± 1·2 | 0·360 |
| Sodium (mg/day) | 1549·6 ± 385·1 | 1568·4 ± 388·9 | 0·815 | 1562·1 ± 391·6 | 1421·1 ± 395·5 | 0·077 | 1434·8 ± 387·9 | 1524·4 ± 391·8 | 0·235 |
| Calcium (mg/day) | 450·4 ± 121·7 | 236·4 ± 101·7 | <0·001* | 409·1 ± 123·8 | 299·7 ± 105·4 | <0·001* | 439·6 ± 122·6 | 244·9 ± 104·4 | <0·001* |
| Iron (mg/day) | 5·2 ± 1·3 | 4·7 ± 1·3 | 0·06 | 5·7 ± 1·3 | 5·1 ± 1·3 | 0·031 | 5·9 ± 1·3 | 4·7 ± 1·3 | <0·001* |
| Zinc (mg/day) | 3·1 ± 0·6 | 2·8 ± 0·8 | 0·094 | 3·5 ± 0·6 | 3·1 ± 0·8 | 0·017 | 3·5 ± 0·7 | 2·6 ± 0·9 | <0·001* |
| Vitamin A, RAE (µg) | 326·0 ± 693·3 | 207·3 ± 106·7 | 0·291 | 386·6 ± 705·1 | 238·7 ± 108·5 | 0·184 | 576·5 ± 698·4 | 174·0 ± 107·5 | <0·001* |
| Beta-carotene (mcg/day) | 449·9 ± 287·4 | 276·9 ± 241·3 | 0·002 | 376·9 ± 292·2 | 377·7 ± 245·4 | 0·988 | 434·1 ± 289·5 | 379·6 ± 243·0 | 0·305 |
| Thiamin (mg/day) | 0·7 ± 0·5 | 0·8 ± 1·1 | 0·922 | 0·9 ± 0·5 | 1·2 ± 1·1 | 0·102 | 0·7 ± 0·5 | 0·8 ± 1·1 | 0·869 |
| Riboflavin (mg/day) | 0·9 ± 0·2 | 0·7 ± 0·3 | <0·001* | 1·1 ± 0·2 | 0·9 ± 0·3 | <0·001* | 1·0 ± 0·2 | 0·7 ± 0·2 | <0·001* |
| Niacin (mg/day) | 7·0 ± 1·9 | 7·0 ± 2·1 | 0·928 | 7·8 ± 2·0 | 8·8± 2·2 | 0·016 | 7·2 ± 2·0 | 7·0 ± 2·2 | 0·485 |
| Vitamin C (mg/day) | 24·3 ± 13·1 | 20·3 ± 12·5 | 0·144 | 19·2 ± 13·3 | 19·1 ± 13·7 | 0·993 | 16·0 ± 13·2 | 17·3 ± 13·6 | 0·612 |

Data are represented as mean ± SD, and significance was determined using repeated measures ANOVA at $P < 0·05$. [a,b]Values in the same row with the same superscript letters are significantly different among the groups at $P < 0·05$. PS=protein substitute with low cholesterol group. WE=whole egg group. RAE= retinol activity equivalents.

**Table S4: Comparison of nutritional intake within groups by day of participants**

| Food categories | Control | PS | WE | *P-value* |
|---|---|---|---|---|
| | | frequency/day/person | | |
| Rice and flour | 0·31[a] | 0·55[b] | 1·69[c] | <0·001* |
| Tuber crop | 0·27[a] | 0·49[b] | 0·18[c] | <0·001* |
| Grains | 0·18[a] | 0·52[a] | 0·03[b] | 0·001* |
| Vegetables | 0·27[a] | 0·63[b] | 0·56[b] | <0·001* |
| Fruits | 0·24[a] | 0·82[b] | 0·17[c] | <0·001* |
| Meat | 0·22[a] | 0·60[b] | 0·95[c] | <0·001* |
| Seafood | 0·25[a] | 0·62[b] | 0·32[c] | 0·001* |
| Eggs | 0·21[a] | 0·40[b] | 0·87[c] | <0·001* |
| Milk | 0·25[a] | 0·54[b] | 0·70[c] | <0·001* |
| Fats | 0·23[a] | 0·70[b] | 0·37[c] | <0·001* |
| Sugar | 0·20[a] | 0·63[b] | 0·30[c] | <0·001* |
| Seasoning | 0·19[a] | 0·50[b] | 0·31[c] | 0·001* |
| Beverages | 0·25[a] | 0·60[b] | 0·53[b] | <0·001* |
| Snack | 0·19[a] | 0·76[b] | 0·16[a] | <0·001* |
| Delicatessen | 0·21[a] | 0·60[b] | 0·51[b] | <0·001* |
| Local food | 0·26[a] | 0·60[b] | 0·72[b] | <0·001* |
| Dessert | 0·21[a] | 0·82[b] | 0·11[c] | <0·001* |
| Soup | 0·23[a] | 0·57[b] | 0·22[a] | <0·001* |

Data are represented as frequency, and significance was determined using repeated measures ANOVA at *P*< 0·05. [a,b]Values in the same row with the same superscript letters are significantly different among the groups at *P*< 0·05. PS=protein substitute with low cholesterol group. WE=whole egg group.

**Table S5: Food pattern of participants**

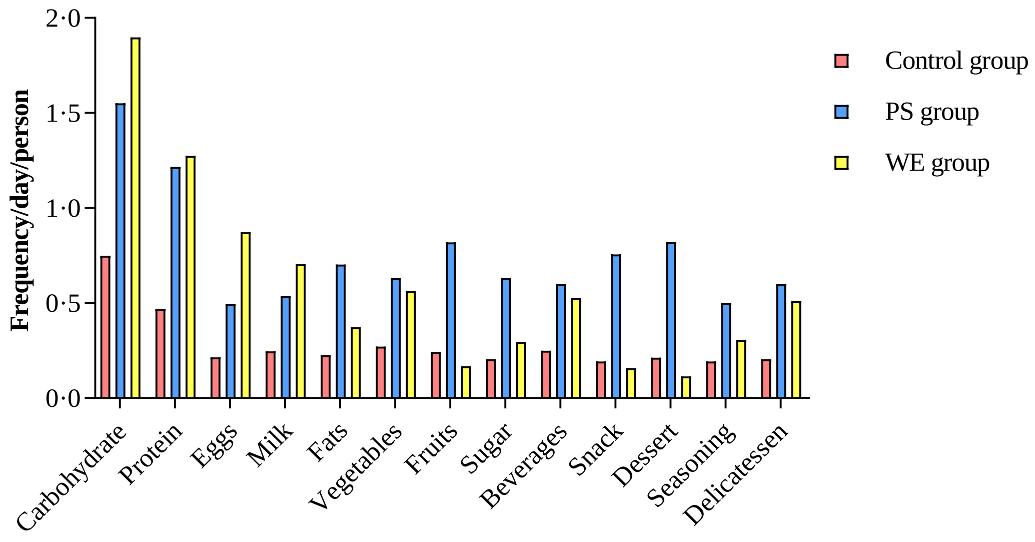


***Figure S1*: Food pattern in study group.**

Bar graph represents frequency per day per person. PS=protein substitute with low cholesterol group. WE=whole egg group.
